# Directional genetic relationships between obsessive-compulsive disorder and bipolar disorder and schizophrenia

**DOI:** 10.64898/2026.05.01.26352245

**Authors:** Maria Niarchou, Marina Natividad Avila, Behrang Mahjani, Joseph D. Buxbaum, Niamh Mullins, Dorothy E. Grice

## Abstract

**Objective:** Obsessive-compulsive disorder (OCD) frequently co-occurs with bipolar disorder (BD) or schizophrenia (SCZ), and, importantly, can often precede their onset. However, the genetic architecture and directionality underlying these relationships remain unclear. We leveraged large-scale genome-wide association study (GWAS) data to examine shared genetic architecture and directional relationships among OCD, BD and SCZ, and used major depressive disorder (MDD) as a comparator.

**Methods:** Using linkage disequilibrium score regression (LDSC), MiXeR, and Generalized Summary-data-based Mendelian Randomization (GSMR) as well as complementary Mendelian randomization approaches, we characterized genetic correlations, polygenic overlap (Dice coefficient), and effect direction concordance (ρ_β_) across disorders.

**Results:** We observed substantial genetic correlations between OCD and BD (r_g_=0.37), BD type 2 (BD2) (r_g_=0.54), and SCZ (r_g_=0.39), with a large proportion of shared causal variants between OCD and both BD (Dice=0.85) and SCZ (Dice=0.84). MiXeR analyses indicated that OCD and BD2 share a smaller proportion of causal variants (Dice=0.57) but there is a high concordance of effect directions amongst these causal variants (ρ_β_=0.96), whereas OCD and MDD showed minimal overlap but strong concordance among shared variants (Dice=0.09, ρ_β_=1). Directional GSMR and complementary TwoSampleMR analyses supported a causal effect of genetic risk to OCD on liability to BD (b=0.20, p=1.5×10□□), SCZ (b=0.52, p=9.5×10□²¹), and MDD (b=0.24, p=1.06×10□□), with little evidence for reverse causal effects.

**Conclusions:** Together, these findings indicate that genetic liability to OCD can represent an early component of transdiagnostic psychiatric risk, with implications for understanding and potentially predicting the emergence of broader psychopathology across the life course.

## Introduction

Obsessive-compulsive disorder (OCD) is a chronic neuropsychiatric disorder characterized by recurrent intrusive obsessions coupled with repetitive, ritualized behaviors. This condition affects nearly 240 million people worldwide, including up to 3% of the population over their lifetime^1,2^. Despite the availability of effective, evidence-based treatments, OCD remains profoundly underdiagnosed and undertreated, with the majority of affected individuals failing to receive appropriate care.

Furthermore, epidemiological data have shown significant comorbidity between OCD and other major psychiatric disorders, including bipolar disorder (BD)^3^, schizophrenia (SCZ)^4^ and major depressive disorder (MDD)^5,6,7^. This comorbidity is typically associated with higher clinical severity and poorer outcomes compared to an isolated OCD diagnosis; for example OCD co-occurring with mood or psychotic disorders is linked to more impairing symptoms and functional disability^6^. First-degree relatives of individuals with OCD also exhibit elevated rates of BD and SCZ^8^, and this risk decreases proportionally with genetic distance, consistent with shared heritability. Longitudinal analyses reveal that OCD typically precedes the onset of BD or SCZ, with median lags of approximately 2-3 years^8,9^. Collectively, these findings indicate substantive cross-disorder overlap, but leave unresolved whether they reflect shared genetic architecture, developmental processes, or directional influences across disorders. Clinically, these co-occurrences have important treatment implications. Antidepressants, commonly prescribed for OCD, can increase the risk of treatment-emergent affective switch in vulnerable individuals, particularly those with bipolar disorder type 1 (BD1). Conversely, certain second-generation antipsychotics have been associated with the emergence or exacerbation of obsessive-compulsive symptoms in patients with SCZ or BD^10^, potentially reflecting serotonergic and frontostriatal circuit mechanisms^11^. Growing evidence suggests that these pharmacological vulnerabilities may arise from shared neurobiological substrates across OCD, BD, and SCZ, underscoring the importance of a biologically informed approach to treatment selection, and the need for a clearer understanding of the biological connections among these disorders^12^.

Despite these observations, the mechanistic nature of the OCD-BD and OCD-SCZ relationships remains uncertain. Two non-mutually exclusive explanations are: (i) shared genetic risk (pleiotropy) underlies their co-occurrence^13,14^, or (ii) causal relationships between OCD and these psychiatric disorders^15^. Distinguishing among these possibilities is essential for clarifying whether observed cross-disorder associations reflect shared etiology, developmental progression, or diagnostic structure. Prior studies have begun to interrogate the genetic relationship between SCZ and OCD using Mendelian randomization (MR) and pleiotropy-informed approaches^16,17^, reporting evidence consistent with shared polygenic architecture. One study reported unidirectional genetic effects from SCZ to OCD^16^, though this was not replicated in a subsequent study^17^. However, these studies were limited by smaller OCD GWAS sample sizes, restricting power to evaluate OCD as an exposure, and were largely focused on SCZ alone. As a result, it remains unclear whether the observed genetic overlap reflects shared biological mechanisms or directional genetic liability across disorders, leaving open key questions about whether OCD represents an early factor for broader psychopathology or a consequence of shared liability.

Here, we leverage the latest GWAS summary statistics for OCD, BD (and its subtypes), SCZ, and MDD to systematically characterize their genetic correlation, polygenic overlap, and directional relationships using a suite of robust MR Methods. We focus primarily on BD and SCZ given their clinical and epidemiological overlap with OCD, while including MDD as a comparator to contextualize these findings within a broader psychiatric framework. By jointly examining multiple disorders with well-powered data, we aim to clarify whether OCD’s genetic relationships with BD and SCZ reflect shared architecture, directional effects, or both, with implications for prognosis, early detection, and treatment stratification.

## Methods

### Data Sources

We used publicly available GWAS summary statistics from large-scale psychiatric consortia (**Table 1**). These datasets include the most recent and well-powered studies of OCD^18^, BD, and its subtypes^19,20^, SCZ^18,21^, and MDD^22^. All analyses were constrained to individuals of European ancestry, as available GWAS summary statistics were predominantly derived from European-ancestry samples; this restriction also minimized confounding due to population structure.

**Table 1:** GWAS summary statistics used in this study.

| **Table 1:** GWAS summary statistics used in this study. |  |  |  |  |  |
| --- | --- | --- | --- | --- | --- |
| **Disorder** | **Author** | **PMID** | **N<sub>case</sub>** | **N<sub>control</sub>** | **Year** |
| OCD | *Ström et al.* | 40360802 | 53,660 | 2,044,417 | 2025 |
| BD–PGC3 | *Mullins et al.* | 34002096 |  |  | 2021 |
| BD |  |  | 41,917 | 371,549 |  |
| BD1 |  |  | 25,060 | 449,978 |  |
| BD2 |  |  | 6,781 | 364,075 |  |
| BD–PGC4 | *O'Connell et al.* | 39843750 |  |  | 2024 |
| BD–Clinical |  |  | 27,196 | 43,792 |  |
| BD–Community |  |  | 32,091 | 737,230 |  |
| SCZ | *Trubetskoy et al.* | 35396580 | 53,386 | 47,258 | 2022 |
| MDD | *Adams et al.* | 39814019 | 26,778 | 51,857 | 2025 |

**Abbreviations:** OCD, obsessive-compulsive disorder; BD, bipolar disorder; BD1, bipolar disorder type 1; BD2, bipolar disorder type 2; SCZ, schizophrenia; MDD, major depressive disorder, s.e., standard error, Dice=Dice coefficient, $\rho_{\beta}$ =SNP effect sizes

For each exposure and outcome, disease status was defined in the source GWAS using standardized diagnostic criteria appropriate to each disorder as explained below. Full details of case definitions, control selection, and diagnostic assessment methods are provided in the original GWAS publications cited in **Table 1**.

#### Obsessive-Compulsive Disorder

OCD cases were defined using clinician-confirmed diagnoses and/or structured diagnostic interviews aligned with DSM-IV or DSM-5 criteria as well as diagnoses derived from electronic health records or biobank data, including ICD-coded diagnoses in some contributing cohorts. In addition, large population-based cohorts, including 23andMe, contributed self-reported OCD diagnoses. Control participants were screened to exclude OCD and other major psychiatric disorders when possible.

#### Bipolar Disorder

We used summary statistics from two major waves of GWAS conducted by the Psychiatric Genomics Consortium (PGC).

The PGC3 bipolar disorder study^19^ provided results for BD overall (henceforth BD-PGC3), as well as subtype-specific analyses of BD type 1 (BD1-PGC3), and BD type 2 (BD2-PGC3). BD cases were defined using standardized criteria, primarily based on DSM-IV, based on structured interviews, clinician assessment, or medical record review. BD1-PGC3 was defined by the presence of manic episodes, whereas BD2-PGC3 required hypomanic episodes and major depressive episodes without a history of full mania. Controls were screened to exclude BD and related psychiatric conditions.

Additionally, we included the more recent PGC4 BD GWAS^20^, which stratified cases into clinically ascertained (BD-clinical-PGC4) and community-based (BD-community-PGC4) cohorts. The clinical subset included individuals diagnosed through psychiatric services using structured interviews or clinician confirmation, while the community subset incorporated population-based cohorts and biobank samples where diagnoses were derived from electronic health records, self-report, or algorithmic phenotyping.

#### Schizophrenia

Schizophrenia summary statistics were obtained from a large-scale meta-analysis conducted by the PGC^21^. Cases were diagnosed according to DSM-IV or ICD-10 criteria using structured clinical interviews, clinician assessment, or medical record review. Controls were screened to exclude psychotic disorders.

#### Major Depressive Disorder

MDD cases were defined in the latest paper of the Major Depressive Disorder Working Group of the Psychiatric Genomics Consortium^22^ using structured diagnostic interviews, clinician diagnosis, or validated questionnaire-based assessments consistent with DSM-IV or DSM-5 criteria. Control participants were screened to exclude lifetime MDD diagnosis.

Analyses were performed using summary-level GWAS data only. Individual-level genotype information was not accessed. This study was reported in accordance with the Strengthening the Reporting of Observational Studies in Epidemiology (STROBE) guidelines for observational studies^23^ (see **Supplement)**. Ethical approval for each contributing GWAS was obtained from the relevant institutional review boards in the original studies, and our analysis was not pre-registered. The present study used de-identified summary data and did not require additional ethical approval.

### Measurement, quality control, and selection of genetic variants

Genetic variants were measured as single nucleotide polymorphisms (SNPs) represented by per-allele effect size estimates and standard errors from GWAS summary statistics. All variants included in the present analyses had undergone quality control, including standard procedures for genotyping, imputation, and variant-level filtering, as described in the respective source publications.

Missing genotype or phenotype data were addressed in the original GWAS through study-specific quality control and imputation procedures, as described in the respective source publications. In the present analyses, only SNPs with complete summary statistics (effect size estimates and standard errors) available for both exposure and outcome datasets were included. No additional imputation or modeling of missing data was performed.

### Genetic Correlation

We used linkage disequilibrium score regression (LDSC)^24^ to estimate the genetic correlation (*r □*) between OCD and each psychiatric disorder. LDSC regresses GWAS effect size products onto LD scores derived from the 1000 Genomes European reference panel, correcting for population stratification and sample overlap.

### Polygenic Architecture: MiXeR

To estimate the degree and structure of shared polygenicity, we applied MiXeR (v1.3)^25,26^ to all OCD-disorder pairs. MiXeR is a causal mixture model that decomposes genome-wide heritability into unique and shared causal variant components, independent of causal direction. MiXeR quantifies the degree of polygenic overlap, using the Dice similarity index, which reflects the proportion of shared causal variants between traits (ranging from 0 to 1). MiXeR also estimates the concordance of effect directions among shared variants (ρ_β_), allowing assessment of whether overlapping genetic influences tend to act in the same or opposite directions across traits.

### Mendelian Randomization

Mendelian randomization uses genetic variants as instrumental variables to estimate the effect of an exposure on an outcome, leveraging the random allocation of alleles at conception to reduce confounding and reverse causation^27^. Causal inference using MR relies on three core assumptions: (i) the genetic instruments are robustly associated with the exposure (relevance); (ii) the instruments are independent of confounders of the exposure–outcome relationship (independence); and (iii) the instruments influence the outcome only through the exposure and not through alternative biological pathways (exclusion restriction)^27^. Violations of the exclusion restriction assumption may occur in the presence of horizontal pleiotropy whereby genetic variants affect the outcome through pathways independent of the exposure.

We employed Generalised Summary-data-based Mendelian Randomisation (GSMR)^28^ implemented in GCTA software (version 1.94.1)^29^ to investigate potential bidirectional causal relationships between genetic liability to OCD and BD, SCZ, and MDD. Genome-wide significant SNPs (p < 5×10*□□*) were selected as instrumental variables (IVs) and pruned for LD (r^2^ < 0.01) based on the 1000 Genomes European reference panel. To mitigate bias for horizontal pleiotropy, the HEIDI-outlier method (p < 0.01) was used to identify and exclude pleiotropic variants. We tested OCD → BD/SCZ/MDD and BD/SCZ/MDD → OCD directions. Only analyses with ≥10 independent IVs were retained consistent with prior recommendations^28^.

All traits were binary, and SNP effects were modeled on the log-odds scale. Accordingly, causal estimates reflect the change in log-odds of the outcome per one-unit increase in genetically predicted liability to the exposure. No additional transformations were applied. Covariate adjustment (e.g., age, sex, ancestry principal components) was performed within the original GWAS from which summary statistics were derived.

To evaluate the robustness of findings and further explore potential violations of these assumptions, we conducted complementary analyses using the TwoSampleMR package (v0.5.6) in R. These sensitivity analyses included inverse-variance weighted (IVW), MR-Egger, weighted median, simple mode, and weighted mode estimators. Between-instrument heterogeneity was quantified using Cochran’s Q statistics under both IVW and MR-Egger models. Directional horizontal pleiotropy was assessed using the MR-Egger intercept test, and outlier-driven pleiotropy was evaluated using MR-PRESSO^30^. Instrument strength was evaluated using the mean F-statistic across all selected SNPs, with values below 10 considered indicative of weak instruments. To assess the reliability of MR-Egger estimates, we calculated the I²_GX_ statistic as a measure of potential regression dilution bias. Directionality was further assessed using Steiger filtering to confirm that instruments explained greater variance in the exposure than in the outcome; variants failing this criterion were considered indicative of potential reverse causation. Concordance in the direction and magnitude of effect estimates across methods was used to support the stability of GSMR findings.

Information regarding participant-level overlap between exposure and outcome GWAS was not available. Although partial sample overlap across psychiatric consortia cannot be ruled out, any such overlap is expected to represent a small fraction of the total sample size and is therefore unlikely to substantially bias effect estimates^31^.

We conducted power calculations for all MR analyses to estimate the minimum detectable effect size at 80% statistical power using the mRnd calculator. These calculations were not used to determine analytic decisions but were performed to aid interpretation of effect estimates^32^.

We did not apply multiple testing correction, as GSMR was specified as the primary analysis. Complementary Mendelian randomization methods were used as sensitivity analyses to assess robustness of findings rather than as independent hypothesis tests.

## Results

### Genetic Correlations

LDSC revealed significant positive genetic correlations across all traits examined (Figure 1, STable 1). The strongest correlation was observed between OCD-MDD (rg = 0.56, p = 3.6×10*□*^3^*□*), followed by OCD-BD2-PGC3 (rg = 0.54, p = 4.7×10*□*^2^*□*), OCD-BD-Community-PGC4 (r_g_ = 0.45, p = 8.71 x 10-30), and OCD-SCZ (r_g_ = 0.39, p = 1.6×10-65) (Figure 1). The correlation with BD1-PGC3 (r_g_ = 0.29, p = 2.2 x 10-26) was markedly weaker than with BD2-PGC3, suggesting subtype-specific genetic relationships.

**Figure 1.**
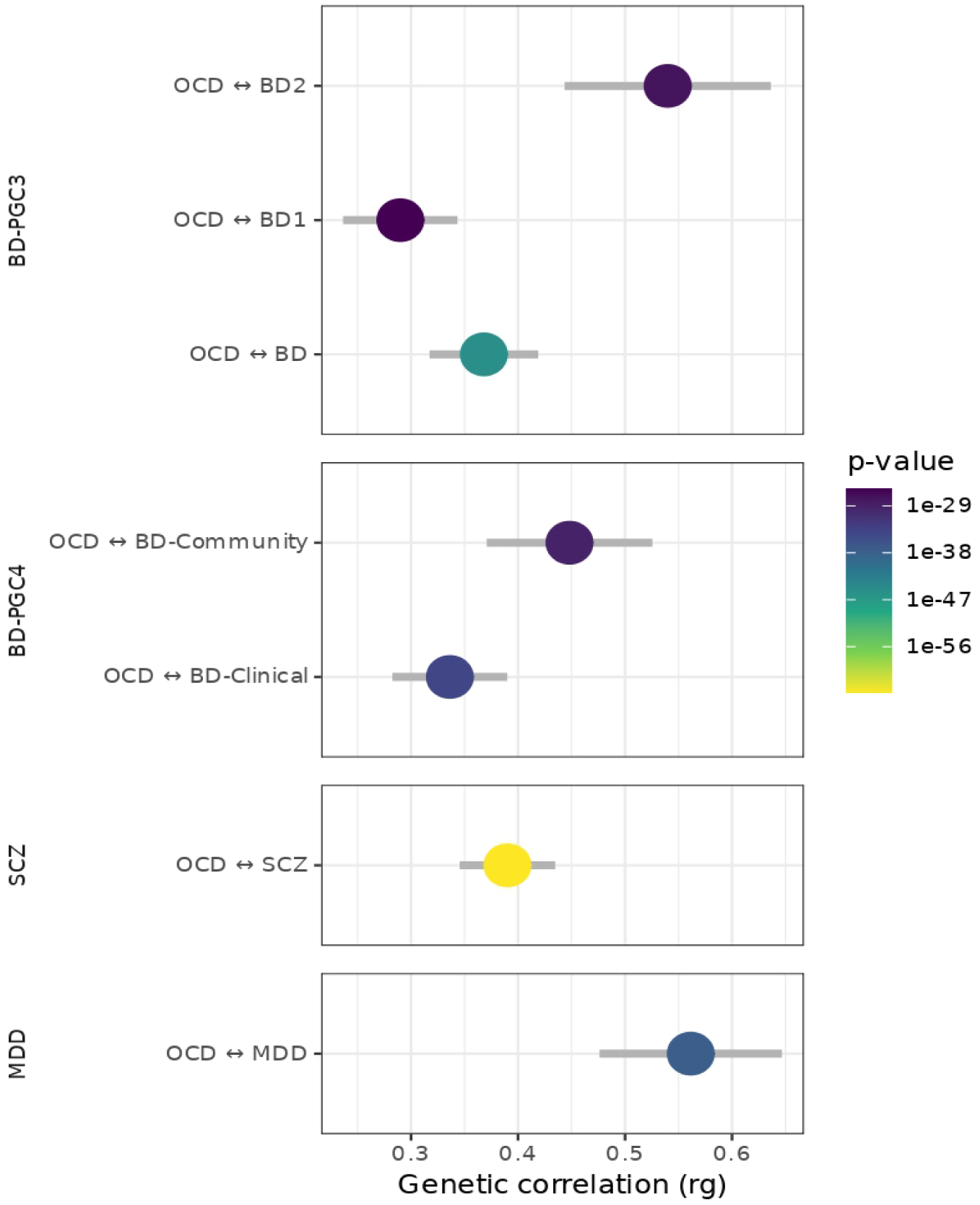
Genetic correlations (r_9_) between obsessive-compulsive disorder (OCD) and bipolar disorder (BD; including BD type 1 (BD1) and type 2 (BD2)), schizophrenia (SCZ), and major depressive disorder (MDD) estimated using linkage disequilibrium score regression (LDSC). Points represent estimated genetic correlations, and horizontal error bars indicate standard errors. Color intensity reflects statistical significance (p-values), with more extreme colors indicating smaller p-values. Results are shown separately for BD-PGC3 (including BD, BD1, BD2), BD-PGC4 (clinical and community subsets), SCZ, and MDD. **Abbreviations:** OCD, obsessive-compulsive disorder; BD, bipolar disorder; BD1, bipolar disorder type 1; BD2, bipolar disorder type 2; SCZ, schizophrenia; MDD, major depressive disorder; r_₉_, genetic correlation.

### Shared Polygenic Architecture

Across BD phenotypes, MiXeR analyses indicated substantial polygenic overlap with OCD (**Figure 2**, **Table 2**). We observed high Dice coefficients for BD-PGC3 (Dice=0.85) and BD1-PGC3 (Dice=0.80), indicating substantial sharing of OCD-associated variants with BD. This overlap was accompanied by moderate concordance in effect direction (OCD-BD1-PGC3 ρ_β_=0.36; OCD-BD2-PGC3 ρ_β_=0.44). The strongest alignment of effect sizes was observed for OCD-BD2-PGC3, which showed lower overall overlap (Dice = 0.57) but very high concordance among the shared variants (ρ_β_ = 0.96), suggesting a more restricted subset of variants with highly aligned effects. Comparisons across clinically ascertained (BD-clinical-PGC4) and community-based (BD-community-PGC4) BD samples revealed broader overlap of OCD with clinical cohorts (Dice = 0.79, ρ_β_ =0.44) but stronger effect concordance with community samples (Dice = 0.46, ρ_β_ = 0.82).

**Figure 2.**
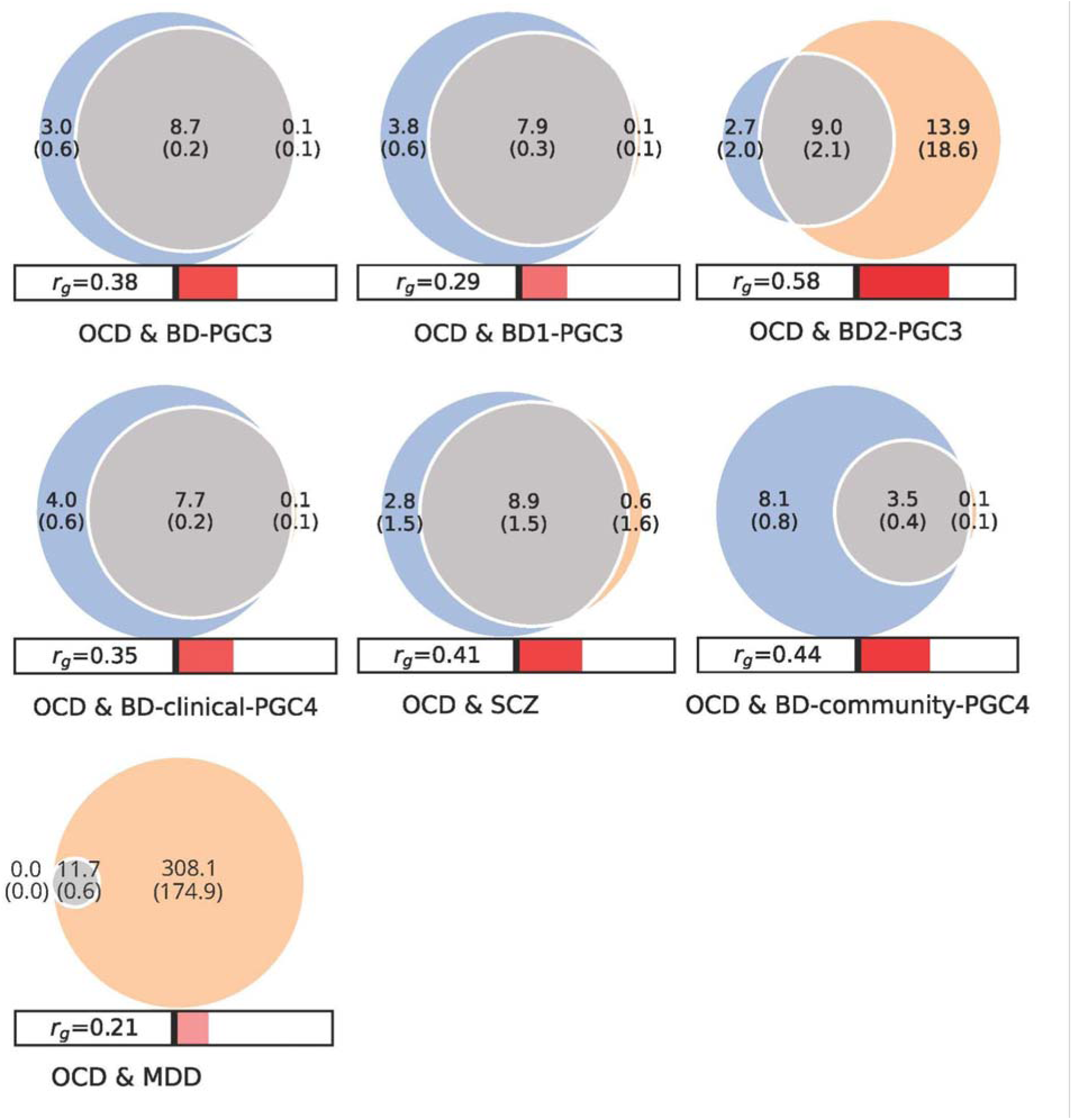
Polygenic overlap between OCD and related psychiatric disorders using MiXeR. Venn diagrams illustrating the estimated number of trait-influencing variants unique to OCD (blue), unique to each comparison disorder (i.e., BD, SCZ, and MDD) (orange), and shared between disorders (grey). Values represent the estimated number of causal variants in thousands with standard errors shown in parentheses. Bars below each diagram indicate the genetic correlation estimated using MiXeR. **Abbreviations:** OCD, obsessive-compulsive disorder; BD, bipolar disorder; BD1, bipolar disorder type 1; BD2, bipolar disorder type 2; SCZ, schizophrenia; MDD, major depressive disorder; r_₉_, genetic correlation.

**Table 2:** Polygenic overlap and effect concordance between OCD and BD/SCZ/MDD.

| | *Dice* | *S.e.*<sub>Dice</sub> | $\rho_{\beta}$ | *S.e.*<sub>$\rho_{\beta}$</sub> |
| --- | --- | --- | --- | --- |
| **BD-PGC3** |  |  |  |  |
| OCD ↔ BD | 0.852 | 0.028 | 0.441 | 0.017 |
| OCD ↔ BD1 | 0.801 | 0.026 | 0.360 | 0.015 |
| OCD ↔ BD2 | 0.569 | 0.092 | 0.960 | 0.048 |
| **BD-PGC4** |  |  |  |  |
| OCD ↔ BD–Clinical | 0.789 | 0.029 | 0.439 | 0.019 |
| OCD ↔ BD–Community | 0.463 | 0.053 | 0.822 | 0.075 |
| **SCZ** |  |  |  |  |
| OCD ↔ SCZ | 0.837 | 0.143 | 0.512 | 0.155 |
| **MDD** |  |  |  |  |
| OCD ↔ MDD | 0.088 | 0.043 | 1.000 | 0.000 |

**Abbreviations:** OCD, obsessive-compulsive disorder; BD, bipolar disorder; BD1, bipolar disorder type 1; BD2, bipolar disorder type 2; SCZ, schizophrenia; MDD, major depressive disorder, s.e., standard error, Dice=Dice coefficient, $\rho_{\beta}$ =SNP effect sizes

For schizophrenia, MiXeR likewise indicated extensive polygenic sharing with OCD (Dice = 0.84), with moderate concordance of effect sizes among shared variants (ρ_β_ = 0.51).

In contrast, OCD-MDD comparisons revealed a different pattern. MiXeR estimated very limited polygenic overlap (Dice = 0.09), indicating that only a small fraction of OCD-associated variants also contributed to MDD liability. However, among this restricted subset of shared variants, effect sizes were highly concordant (ρ_β_ = 1.00), indicating that the genetic components linking OCD and MDD are concentrated in a narrow but strongly aligned set of loci.

### Mendelian Randomization

#### Instrument strength and statistical power

Power calculations indicated that we had 80% power to detect small effect sizes (OR 1.02 to 1.14) across all analyses (**STables 2- 13**). All instruments met standard strength criteria (mean F > 10; **STables 2-13**), supporting the reliability of the MR analyses.

#### GSMR

Bidirectional GSMR analyses indicated asymmetric genetic relationships between OCD and the comparison disorders (**Table 3, STables 2-12**). When OCD was modeled as the outcome, none of the BD or SCZ GWAS showed statistically significant effects on OCD, with all estimates small and non-significant (all p > 0.09), providing no evidence of causal effects of BD or SCZ on OCD. Individual SNP associations are summarized through aggregate MR estimates, with full results provided in **Table 4** and **STables 2 to 5**.

**Table 3:** Bidirectional GSMR analyses between OCD and BD/SCZ/MDD.

| Outcome OCD |  |  |  |  | Exposure OCD |  |  |  |  |
| --- | --- | --- | --- | --- | --- | --- | --- | --- | --- |
| Exposure | *b* | *s.e.* | *p* | <sup>N</sup>SNP | Outcome | *b* | *s.e.* | *p* | <sup>N</sup>SNP |
| BD–PGC3 | 0.0331 | 0.0243 | 0.173164 | 23 | BD–PGC3 | **0.2007** | **0.0529** | 1.50E-04 | **20** |
| BD1–PGC3 | 0.0292 | 0.0172 | 0.090013 | 30 | BD1–PGC3 | **0.2226** | **0.0630** | 4.10E-04 | **21** |
| BD2–PGC3 | n/a | n/a | n/a | n/a | BD2–PGC3 | **0.4102** | **0.1048** | 9.03E-05 | **22** |
| BD–clinical–PGC4 | 0.0098 | 0.0213 | 0.647 | 24 | BD–clinical–PGC4 | **0.1713** | **0.0576** | 2.93E-03 | **20** |
| BD–community–PGC4 | n/a | n/a | n/a | n/a | BD–community–PGC4 | **0.2189** | **0.0578** | 1.54E-04 | **22** |
| SCZ | 0.0320 | 0.0191 | 0.093843 | 37 | SCZ | **0.5224** | **0.0559** | 9.54E-21 | **18** |
| MDD | n/a | n/a | n/a | n/a | MDD | **0.2391** | **0.0617** | 1.06E-04 | **22** |

**Abbreviations:** BD, bipolar disorder; BD1, bipolar disorder type 1; BD2, bipolar disorder type 2; SCZ, schizophrenia; MDD, major depressive disorder.

In contrast, when OCD was modeled as the exposure, strong and consistent effects were observed on all outcomes using GSMR (**Table 3**). A one-unit increase in genetically predicted liability to OCD (on the log-odds scale) was associated with increased risk of BD across all definitions, including BD-PGC3 (b = 0.20, s.e. = 0.05, p = 1.5 × 10*□□*), BD1-PGC3 (b = 0.22, s.e. = 0.06, p = 4.1 × 10*□□*), BD2-PGC3 (b = 0.41, s.e. = 0.10, p = 9.0 × 10*□□*), BD-clinical-PGC4 (b = 0.17, s.e. = 0.06, p = 2.9 × 10*□*^3^), and BD-community-PGC4 (b = 0.22, s.e. = 0.06, p = 1.5 × 10*□□*). OCD liability was also strongly associated with SCZ (b = 0.52, s.e. = 0.06, p = 9.5 × 10*□*^21^) and with MDD (b = 0.24, s.e. = 0.06, p = 1.1 × 10*□□*).

#### Sensitivity analyses

When OCD was modeled as the outcome, sensitivity analyses using TwoSampleMR were broadly consistent with the primary GSMR findings (**STables 2-5**). Across BD phenotypes, effect estimates were small and non-significant across IVW and weighted median models, with no consistent evidence of directional effects across mode-based and MR-Egger results. For SCZ, weighted median and MR-Egger results showed nominal significance; however, effect estimates were inconsistent across methods. Although heterogeneity was observed for BD1-PGC3 and BD-clinical-PGC4, MR-Egger intercept tests and MR-PRESSO analyses did not indicate substantial directional pleiotropy, and exclusion of identified outliers did not materially alter the causal estimates. Steiger filtering supported the hypothesized causal direction when OCD was modeled as the outcome, with approximately 100% of variants explaining greater variance in the exposure than in OCD across all traits (**STables 2-5**).

When OCD was modeled as the exposure, sensitivity analyses were consistent with the GSMR findings across all outcomes (**STables 6-13**). For BD phenotypes, IVW and weighted median estimates were directionally concordant with GSMR, with largely overlapping effect sizes; MR-Egger estimates were also consistent except for BD-community-PGC4, where the effect direction differed. Cochran’s Q statistics indicated heterogeneity in most BD analyses, whereas MR-Egger intercept tests were generally non-significant, with the exception of BD-community-PGC4. MR-PRESSO identified evidence of horizontal pleiotropy for BD-PGC3, BD1-PGC3, and BD-community-PGC4; however, distortion tests were not significant, and exclusion of identified outliers did not substantially alter effect estimates (**STable 13**). For instance, for OCD➔BD1-PGC3, removal of two outlier variants attenuated the GSMR estimate (0.22 to 0.14) but the association remained statistically significant. Also, When OCD was modeled as the exposure, Steiger filtering indicated strong support for the assumed causal direction across most bipolar disorder phenotypes (90–100% of variants explaining greater variance in OCD than in the outcome), although support was weaker for BD–clinical (55%).

For the effect of OCD on SCZ, all MR methods yielded overlapping effect size estimates and consistent directions of effect relative to the GSMR (**STable 11**). Cochran’s Q statistics were significant, but the MR-Egger intercept test showed no evidence of directional pleiotropy. Similarly, although the MR-PRESSO global test showed evidence of horizontal pleiotropy and identified 3 outliers, the distortion test was not significant. Steiger filtering provided limited support for the assumed causal direction, with 33% of variants explaining greater variance in OCD than in schizophrenia.

Modeling OCD as an exposure on MDD, all MR methods yielded overlapping effect size estimates and consistent directions of effect relative to the GSMR (**STable 12**). Neither Cochran’s Q statistics nor the MR-Egger intercept test were significant. Similarly, the MR-PRESSO global test did not provide evidence for horizontal pleiotropy. Steiger filtering showed that 73% of variants explained greater variance in OCD than in MDD, indicating moderate support for the assumed causal direction.

## Discussion

Our study provides the first comprehensive evaluation of shared genetic architecture and bidirectional causal relationships between OCD and other major psychiatric conditions, namely SCZ, BD, and MDD. The possibility that risk for a subsequent BD or SCZ diagnosis in an individual with OCD might be assessed before BD or SCZ symptom emergence provided the strong motivation for exploring these relationships further. We added MDD as a comparator because it can also co-occur with OCD, an OCD diagnosis can precede an MDD diagnosis, and, if severe, treatment resistant or associated with psychotic features, MDD represents a severe mental illness.

Beyond confirming substantial genetic correlations between OCD and each of these disorders, consistent with prior work^33^, we quantify the degree of polygenic overlap and provide convergent evidence supporting directional genetic effects from OCD liability to BD, SCZ, and MDD. Together, these findings refine our understanding of cross-disorder genetic architecture and support a model in which OCD liability represents an upstream component of transdiagnostic psychiatric risk.

MiXeR analyses indicated that OCD shares a large proportion of its polygenic signal with BD and SCZ, consistent with substantial polygenic overlap across genetic variants. In contrast, overlap with MDD was substantially smaller, reflecting largely disorder-specific polygenic components. However, the shared variants between OCD and MDD demonstrated near complete concordance in effect sign, indicating that although fewer variants are shared, their effects are highly aligned. These findings highlight both shared and disorder-specific components of the genetic architecture linking OCD with these other psychiatric phenotypes, with differing degrees and patterns of overlap.

Directional GSMR analyses further refined this picture by revealing a pronounced asymmetry in genetic relationships across disorders. When OCD was modeled as the outcome, genetic liability to BD or SCZ showed little evidence of association with OCD. In contrast, when OCD was modeled as the exposure, strong and consistent associations were observed across all outcomes, including multiple definitions of BD as well as SCZ and MDD. This pattern was supported by complementary Mendelian randomization analyses, which showed concordant effect directions and similar effect sizes across inverse-variance weighted and weighted median estimators, with no evidence of directional horizontal pleiotropy. The consistency across analytic approaches and the application of HEIDI-outlier filtering reduce the likelihood that these findings are driven by pleiotropic variants or instrument instability. Steiger filtering generally supported the hypothesized direction of effect for bipolar disorder phenotypes and MDD, although support was weaker for schizophrenia. Collectively, these findings suggest that genetic liability to OCD is causally associated with liability to other major psychiatric disorders, whereas the reverse pattern is not observed, highlighting OCD as a potentially upstream component within the shared genetic architecture of severe mental illness.

### Potential biological mechanisms

The observed directional genetic associations may reflect shared neurobiological pathways through which liability to OCD contributes to broader psychiatric vulnerability. OCD has been linked to alterations in cortico-striatal-thalamo-cortical circuitry^34^, glutamatergic and serotonergic signaling^35^, and neurodevelopmental processes^36^ that overlap with pathways implicated in BD, SCZ, and MDD. Emerging genetic and transcriptomic evidence^37–39^ suggests that OCD risk is enriched in genes involved in chromatin regulation, pointing to upstream mechanisms that influence gene expression during brain development and in response to environmental factors^40^. Notably, OCD typically has an earlier age of onset than bipolar or psychotic disorders, with approximately 25% of cases emerging by age 14, 50% by age 19, and nearly two-thirds by age 25^1^. This raises the possibility that genetic liability to OCD may influence early neurodevelopmental trajectories or circuit maturation in ways that increase susceptibility to later-onset psychiatric conditions. This temporal pattern is consistent with the asymmetric directionality observed in the present analyses. ‘

The especially strong association between OCD and BD2 may further reflect overlapping neurobiological substrates related to mood regulation^41,42^, reward sensitivity^43–45^, and cognitive control^46,47^. BD2 is characterized by recurrent depressive and hypomanic episodes, and is frequently associated with persistent depressive burden, significant functional impairment and emotional dysregulation^48^. Meta-analytic evidence also supports consistent deficits in cognitive flexibility and executive control in OCD, domains that may also be affected in BD2^48^. These shared features suggest that BD2 and OCD may share underlying processes that could reflect chronicity, and cognitive inflexibility- features that show conceptual and neurobiological overlap with OCD. Shared dysregulation of fronto-striatal circuits^49–53^ and glutamatergic signaling^54,55^ may therefore contribute to the pronounced genetic coherence observed between these conditions.

Finally, it must be considered that in some cases, emergent BD or SCZ may represent a later manifestation of what first appears as OCD, rather than an entirely separate condition; a process termed pseudo-comorbidity by Caron and Rutter (1991)^56^. Caron and Rutter discuss other relevant mechanisms of apparent comorbidity, all of which are of potential interest here. These include disorders reflecting extremes on shared dimensions rather than distinct entities, overlapping diagnostic criteria or artificial subdivision inflating apparent co-occurrence, or one disorder being part of the other.

### Clinical implications

These findings may have clinical relevance for understanding patterns of psychiatric comorbidity. OCD frequently co-occurs with BD and SCZ, and such co-occurrences have an important impact on treatment approaches and clinical outcomes. Clarifying the biology of these disorders may help determine whether comorbid presentations represent shared etiological mechanisms, developmental progression or distinct co-occurring conditions, which in turn has implications for treatment paths. While Mendelian randomization estimates reflect the effects of lifelong genetic liability rather than the consequences of clinical diagnosis or treatment, the observed directional genetic associations suggest that liability to OCD may mark individuals at increased risk for broader psychiatric vulnerability. From a clinical perspective, this supports the importance of careful longitudinal assessment and monitoring for emerging mood or psychotic symptoms in individuals with OCD, particularly those with additional risk factors. However, the present findings do not inform the expected magnitude of benefit from treating OCD on the risk of other psychiatric disorders and should not be interpreted as evidence that intervention on OCD will prevent subsequent illness. Rather, they motivate future longitudinal and interventional studies to evaluate how genetic liability, clinical course, and treatment exposures influence development of other psychiatric disorders and clinical prognosis.

### Strengths and limitations

To our knowledge, this is the first Mendelian randomization investigation examining OCD in relation to multiple major psychiatric disorders, providing novel insight into shared genetic liability across diagnostic boundaries. We employed a rigorous analytical framework using complementary MR methods alongside extensive sensitivity analyses to assess the robustness of findings and potential violations of underlying assumptions. Analyses adhered to STROBE-MR reporting guidelines, enhancing transparency and reproducibility. In addition, we leveraged the largest and most recent genome-wide association studies available, maximizing statistical power and instrument strength. These results, however, should also be interpreted cautiously given the reliance on summary-level data, the possibility of residual pleiotropy despite extensive sensitivity analyses, and limited power for some reverse-direction tests. Moreover, MR estimates reflect effects on latent liability rather than clinical diagnosis.

Causal interpretation in Mendelian randomization depends on the validity of underlying instrumental variable assumptions^57,58^. MR estimates rely on the assumption that genetic liability to a disorder operates through the same biological pathways as the disorder itself. In psychiatry, where environmental exposures and developmental factors strongly shape clinical presentation, this equivalence may be imperfect^57,58^. Accordingly, the present findings are best interpreted as evidence of directional genetic influence rather than direct clinical causation, underscoring the need for future studies integrating genetic data with longitudinal phenotyping, environmental exposures, and neurobiological measures to clarify underlying mechanisms.

Evidence of heterogeneity across genetic instruments was observed in some analyses, as indicated by significant Cochran’s Q statistics. Such heterogeneity is not uncommon in MR studies of complex traits, and does not, in itself, imply directional horizontal pleiotropy^59^. Consistency across sensitivity analyses supports the robustness of the findings. Several additional limitations warrant consideration. First, all analyses were constrained to individuals of European ancestry, reflecting the current composition of available large-scale psychiatric GWAS. This Eurocentric imbalance limits the generalizability of our findings to non-European populations, given the well-established differences in genetic architecture across ancestries^60^. Second, the limited number of genome-wide significant instruments for some BD subtypes reduced statistical power for reverse-direction Mendelian randomization, potentially biasing those estimates toward the null. Third, partial sample overlap between large psychiatric consortia cannot be ruled out, and could bias estimates toward observational associations; however, the use of robust instruments and consistent findings across multiple MR methods mitigate this concern. Finally, heterogeneity in phenotype definitions across contributing GWAS may introduce noise, although the consistency of results across disorders and disorder subgroups supports the robustness of the primary findings.

## Conclusion

OCD shares a substantial proportion of its genetic risk with BD and SCZ, with evidence of asymmetric directionality in genetic liability. In contrast, reverse-direction analyses provided little evidence that genetic liability to BD or SCZ increases risk for OCD. OCD also showed directional associations with MDD. Together with prior studies demonstrating extensive shared genetic architecture across these psychiatric disorders, our findings extend genetic correlation work by suggesting that cross-disorder relationships may be directionally structured rather than purely reciprocal. Our results suggest that genetic liability to OCD may contribute to risk for other severe psychiatric disorders, underscoring the importance of considering directionality in cross-disorder genetic studies. Future work integrating multi-ancestry samples, refined phenotyping, and longitudinal designs will be essential to clarify underlying mechanisms and evaluate potential preventive implications. Of greatest interest would be whether risk for a subsequent diagnosis of BD, SCZ or severe MDD in an individual with OCD might be predicted before symptom emergence and provide opportunities for enhanced care.

## Supporting information

Strobe_Checklist

## Data Availability

The study used already publicly available summary statistics. Statistical code used to perform the analyses is publicly available on GitHub at https://github.com/buxbaum-lab/OCD-BD

## Author Contributions

N.M., B.M., J.D.B. and D.E.G. conceived and designed the study. M.N.A. performed data analysis. D.E.G., N.M., J.D.B., B.M., M.N.A. and M.N. drafted and revised the manuscript. All authors reviewed and approved the final version of the manuscript. D.E.G. supervised the study.

