## Supplementary_Information for "Directional genetic relationships between obsessive-compulsive disorder and bipolar disorder and schizophrenia"

This report includes, for each disorder pair, **one table** (single-SNP analysis), and **three plots** (scatter, forest, and funnel).

### OCD → BD-PGC3

Table 1. MR-PRESSO Single SNP analysis results for OCD → BD-PGC3

| SNP | b | se | p |
| --- | --- | --- | --- |
| rs10877425 | 0.0598 | 0.1955 | 0.7598 |
| rs11125759 | 0.3517 | 0.2362 | 0.1365 |
| rs11263940 | 0.4779 | 0.2367 | 0.0435 |
| rs11768238 | -0.0983 | 0.2433 | 0.6860 |
| rs1555466 | -0.0668 | 0.2364 | 0.7774 |
| rs203768 | 0.8094 | 0.2425 | 8.43e-04 |
| rs2198140 | 0.2553 | 0.2220 | 0.2501 |
| rs34320 | -0.1166 | 0.2357 | 0.6209 |
| rs3899258 | 0.2339 | 0.2278 | 0.3045 |
| rs4129585 | 0.7262 | 0.2127 | 6.38e-04 |
| rs4831130 | -0.1243 | 0.2583 | 0.6304 |
| rs4931 | 0.4887 | 0.2632 | 0.0633 |
| rs6474628 | 0.7582 | 0.2554 | 0.0030 |
| rs6660196 | -0.3062 | 0.2456 | 0.2124 |
| rs67839857 | -0.4927 | 0.2464 | 0.0455 |
| rs7626445 | 0.2963 | 0.2070 | 0.1524 |
| rs78587207 | 0.0347 | 0.2041 | 0.8649 |
| rs8003822 | 0.3334 | 0.2366 | 0.1588 |
| rs9479138 | 0.3959 | 0.2545 | 0.1198 |
| rs9886111 | -0.0366 | 0.2488 | 0.8832 |
| All - Inverse variance weighted | 0.2005 | 0.0773 | 0.0095 |
| All - MR Egger | 0.3130 | 0.8933 | 0.7301 |

**Scatter plot**

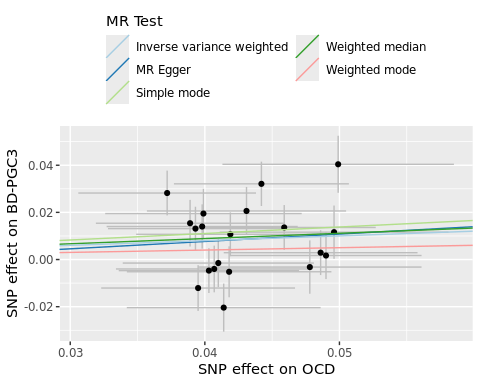

**Forest plot**

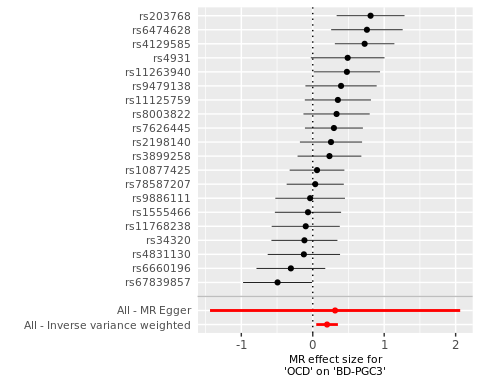

**Funnel plot**

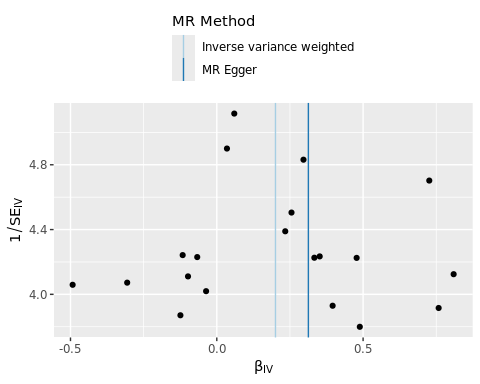

### OCD → BD1-PGC3

Table 2. MR-PRESSO Single SNP analysis results for OCD → BD1-PGC3

| SNP | b | se | p |
| --- | --- | --- | --- |
| rs10877425 | -0.0226 | 0.2448 | 0.9264 |
| rs11125759 | 0.1507 | 0.2914 | 0.6051 |
| rs11263940 | 0.7936 | 0.2901 | 0.0062 |
| rs11768238 | -0.0885 | 0.3022 | 0.7695 |
| rs1555466 | 0.0543 | 0.2908 | 0.8518 |
| rs203768 | 1.1202 | 0.3006 | 1.94e-04 |
| rs2198140 | 0.2577 | 0.2721 | 0.3435 |
| rs34320 | 0.0323 | 0.2903 | 0.9115 |
| rs3899258 | 0.0242 | 0.2802 | 0.9312 |
| rs4129585 | 0.4320 | 0.2624 | 0.0997 |
| rs4702 | 1.3226 | 0.2955 | 7.64e-06 |
| rs4831130 | -0.2129 | 0.3182 | 0.5034 |
| rs4931 | 0.5190 | 0.3233 | 0.1085 |
| rs6474628 | 0.7875 | 0.3145 | 0.0123 |
| rs6660196 | -0.5190 | 0.3038 | 0.0876 |
| rs67839857 | -0.5339 | 0.3019 | 0.0770 |
| rs7626445 | 0.2222 | 0.2615 | 0.3954 |
| rs78587207 | -0.0020 | 0.2531 | 0.9936 |
| rs8003822 | 0.0025 | 0.2926 | 0.9931 |
| rs9479138 | 0.7068 | 0.3136 | 0.0242 |
| rs9886111 | -0.1999 | 0.3073 | 0.5154 |
| All - Inverse variance weighted | 0.2226 | 0.1039 | 0.0322 |
| All - MR Egger | 0.0568 | 1.2195 | 0.9633 |

**Scatter plot**

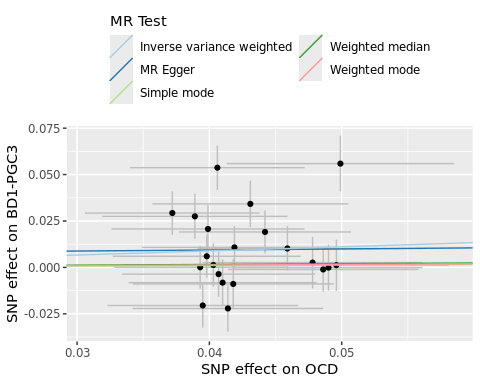

**Forest plot**

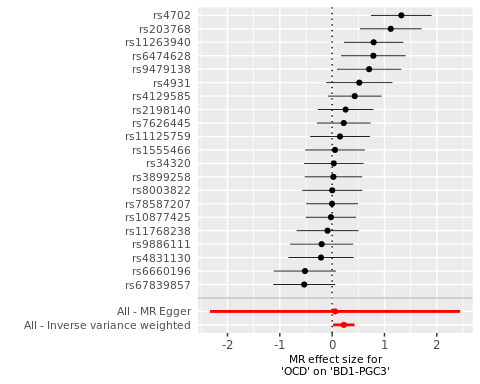

**Funnel plot**

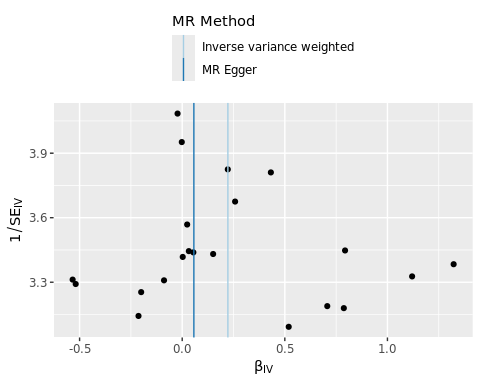

### OCD → BD2-PGC3

Table 3. MR-PRESSO Single SNP analysis results for OCD → BD2-PGC3

| SNP | b | se | p |
| --- | --- | --- | --- |
| rs10877425 | 0.2859 | 0.4197 | 0.4958 |
| rs11125759 | 0.3041 | 0.4950 | 0.5390 |
| rs11263940 | 0.6566 | 0.4966 | 0.1861 |
| rs11768238 | 0.6241 | 0.5160 | 0.2265 |
| rs1555466 | -0.3745 | 0.4937 | 0.4482 |
| rs203768 | 0.6112 | 0.5130 | 0.2335 |
| rs2198140 | 0.4297 | 0.4678 | 0.3584 |
| rs2564930 | 1.0794 | 0.4451 | 0.0153 |
| rs34320 | -0.0446 | 0.4913 | 0.9276 |
| rs3899258 | 0.2481 | 0.4758 | 0.6021 |
| rs4129585 | 1.0135 | 0.4457 | 0.0230 |
| rs4702 | 1.8815 | 0.4950 | 1.44e-04 |
| rs4831130 | -0.3061 | 0.5406 | 0.5712 |
| rs4931 | -0.2231 | 0.5489 | 0.6845 |
| rs6474628 | 1.2851 | 0.5296 | 0.0152 |
| rs6660196 | -0.2710 | 0.5114 | 0.5962 |
| rs67839857 | -0.5724 | 0.5121 | 0.2636 |
| rs7626445 | 0.4641 | 0.4445 | 0.2964 |
| rs78587207 | 0.2857 | 0.4286 | 0.5051 |
| rs8003822 | 0.6667 | 0.4962 | 0.1790 |
| rs9479138 | -0.0798 | 0.5295 | 0.8802 |
| rs9886111 | 0.6148 | 0.5195 | 0.2367 |
| All - Inverse variance weighted | 0.4100 | 0.1238 | 9.27e-04 |
| All - MR Egger | 0.1416 | 1.4542 | 0.9234 |

**Scatter plot**

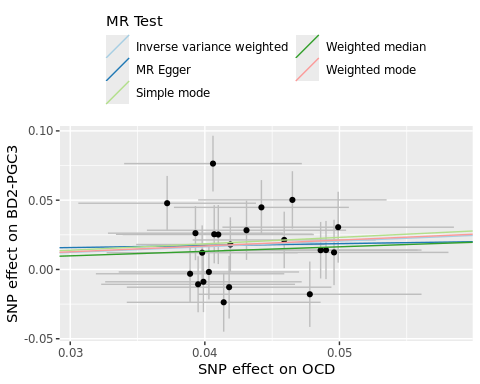

**Forest plot**

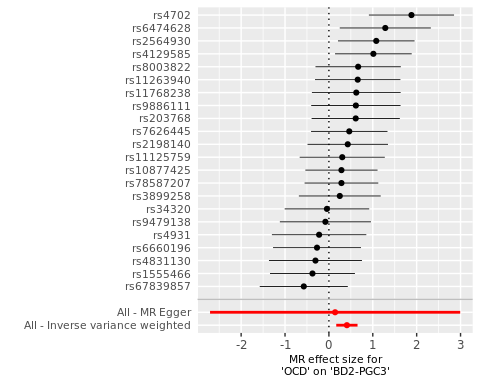

**Funnel plot**

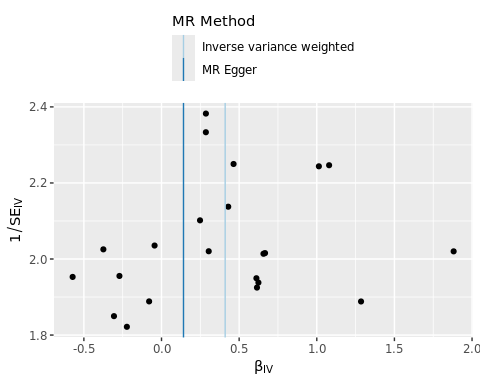

### OCD → BD-clinical-PGC4

Table 4. MR-PRESSO Single SNP analysis results for OCD → BD-clinical-PGC4

| SNP | b | se | p |
| --- | --- | --- | --- |
| rs10877425 | -0.0596 | 0.2119 | 0.7786 |
| rs11125759 | 0.0327 | 0.2613 | 0.9005 |
| rs11263940 | 0.5499 | 0.2576 | 0.0328 |
| rs11768238 | -0.0049 | 0.2703 | 0.9855 |
| rs1555466 | 0.0983 | 0.2615 | 0.7070 |
| rs203768 | 0.7794 | 0.2685 | 0.0037 |
| rs2198140 | 0.3247 | 0.2458 | 0.1866 |
| rs34320 | 0.0472 | 0.2581 | 0.8549 |
| rs3899258 | 0.1452 | 0.2500 | 0.5613 |
| rs4129585 | 0.4728 | 0.2330 | 0.0424 |
| rs4831130 | -0.1531 | 0.2847 | 0.5907 |
| rs4931 | 0.4211 | 0.2883 | 0.1441 |
| rs6474628 | 0.5914 | 0.2796 | 0.0344 |
| rs6660196 | -0.4405 | 0.2709 | 0.1040 |
| rs67839857 | -0.4784 | 0.2681 | 0.0744 |
| rs7626445 | 0.4139 | 0.2331 | 0.0758 |
| rs78587207 | 0.0204 | 0.2265 | 0.9282 |
| rs8003822 | 0.2391 | 0.2621 | 0.3617 |
| rs9479138 | 0.4293 | 0.2802 | 0.1255 |
| rs9886111 | 0.0146 | 0.2732 | 0.9573 |
| All - Inverse variance weighted | 0.1713 | 0.0717 | 0.0168 |
| All - MR Egger | 0.4399 | 0.8257 | 0.6007 |

**Scatter plot**

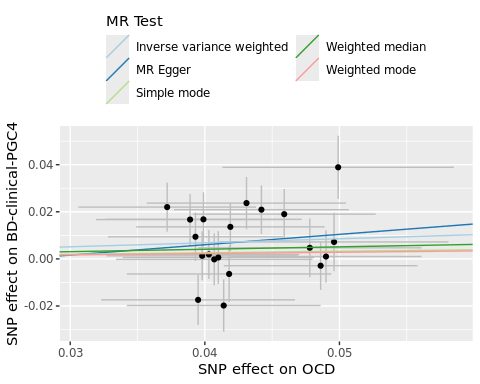

**Forest plot**

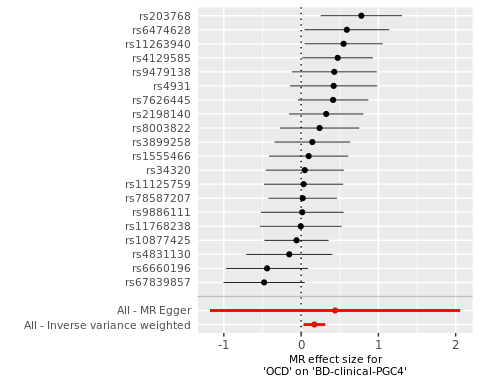

**Funnel plot**

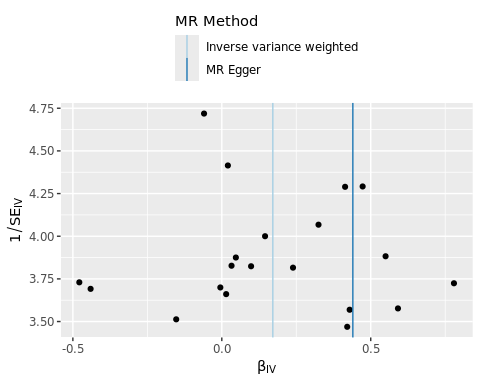

### OCD → BD-community-PGC4

Table 5. MR-PRESSO Single SNP analysis results for OCD → BD-community-PGC4

| SNP | b | se | p |
| --- | --- | --- | --- |
| rs10877425 | -0.1050 | 0.2634 | 0.6901 |
| rs11125759 | 0.6106 | 0.2713 | 0.0244 |
| rs11263940 | 0.1531 | 0.2715 | 0.5728 |
| rs11768238 | 0.3537 | 0.2998 | 0.2380 |
| rs1555466 | -0.2344 | 0.2636 | 0.3740 |
| rs203768 | 0.1523 | 0.2685 | 0.5705 |
| rs2198140 | 0.0836 | 0.2793 | 0.7646 |
| rs2564930 | 0.4667 | 0.2430 | 0.0548 |
| rs34320 | -0.3102 | 0.2754 | 0.2601 |
| rs3899258 | -0.1834 | 0.2722 | 0.5003 |
| rs4129585 | 0.3076 | 0.2421 | 0.2038 |
| rs4702 | 0.8299 | 0.2734 | 0.0024 |
| rs4831130 | 0.1435 | 0.2871 | 0.6172 |
| rs4931 | 0.7119 | 0.3058 | 0.0199 |
| rs6474628 | 0.7043 | 0.2903 | 0.0153 |
| rs6660196 | 0.0936 | 0.2810 | 0.7391 |
| rs67839857 | 0.0605 | 0.2874 | 0.8334 |
| rs7626445 | 0.0872 | 0.2375 | 0.7135 |
| rs78587207 | -0.1979 | 0.2306 | 0.3908 |
| rs8003822 | 0.4249 | 0.2722 | 0.1186 |
| rs9479138 | 0.9381 | 0.2879 | 0.0011 |
| rs9886111 | 0.0976 | 0.2878 | 0.7345 |
| All - Inverse variance weighted | 0.2189 | 0.0751 | 0.0036 |
| All - MR Egger | -2.1434 | 0.7050 | 0.0065 |

**Scatter plot**

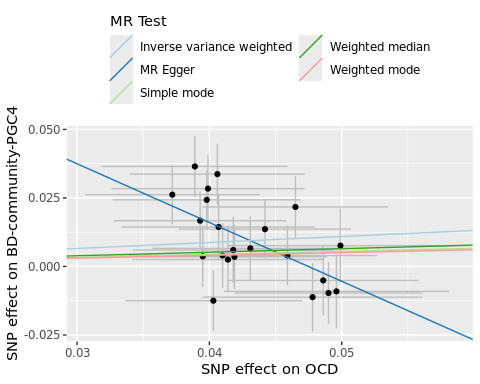

**Forest plot**

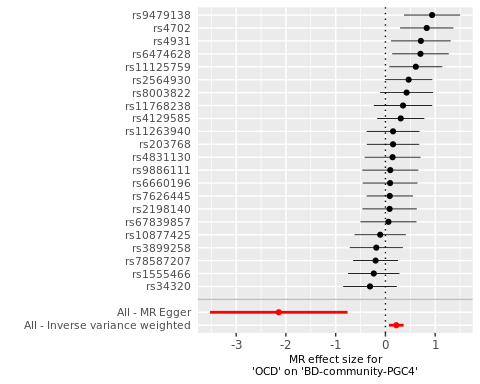

**Funnel plot**

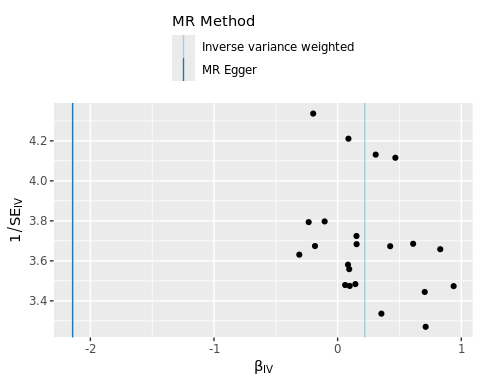

### OCD → SCZ

Table 6. MR-PRESSO Single SNP analysis results for OCD → SCZ

| SNP | b | se | p |
| --- | --- | --- | --- |
| rs11125759 | 0.8618 | 0.2186 | 8.06e-05 |
| rs11263940 | 0.7309 | 0.2158 | 7.07e-04 |
| rs11768238 | 0.2874 | 0.2261 | 0.2035 |
| rs1555466 | 0.3327 | 0.2197 | 0.1299 |
| rs203768 | 1.6312 | 0.2264 | 5.85e-13 |
| rs2198140 | -0.1933 | 0.2053 | 0.3464 |
| rs34320 | 0.5309 | 0.2159 | 0.0139 |
| rs3899258 | -0.1412 | 0.2077 | 0.4965 |
| rs4831130 | 0.4760 | 0.2368 | 0.0445 |
| rs4931 | 1.0127 | 0.2431 | 3.11e-05 |
| rs6474628 | 0.6076 | 0.2339 | 0.0094 |
| rs6660196 | 0.0076 | 0.2279 | 0.9734 |
| rs67839857 | 0.5242 | 0.2246 | 0.0196 |
| rs7626445 | 0.6842 | 0.1961 | 4.85e-04 |
| rs78587207 | 0.9550 | 0.1878 | 3.64e-07 |
| rs8003822 | 0.5368 | 0.2188 | 0.0142 |
| rs9479138 | -0.1157 | 0.2339 | 0.6209 |
| rs9886111 | 0.6513 | 0.2293 | 0.0045 |
| All - Inverse variance weighted | 0.5203 | 0.1081 | 1.50e-06 |
| All - MR Egger | 1.7238 | 1.2343 | 0.1816 |

**Scatter plot**

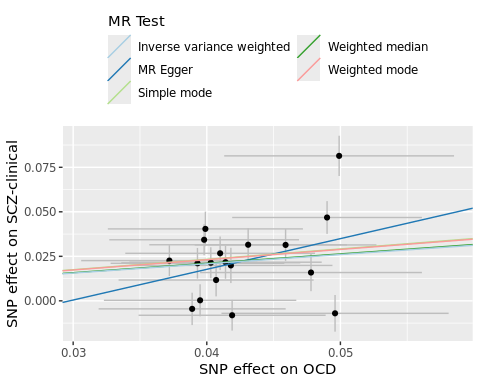

**Forest plot**

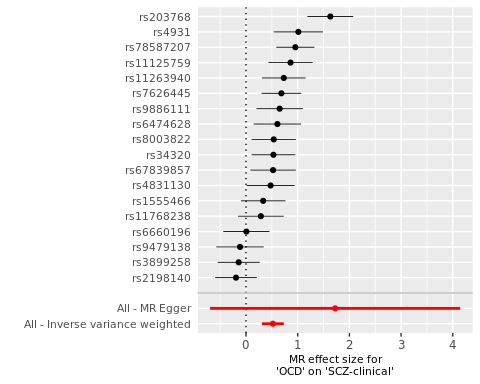

**Funnel plot**

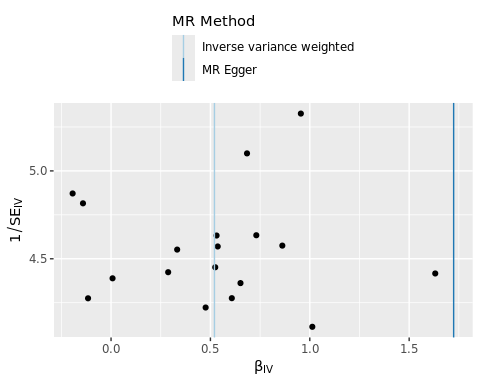

### OCD → MDD

Table 7. MR-PRESSO Single SNP analysis results for OCD → MDD

| SNP | b | se | p |
| --- | --- | --- | --- |
| rs10877425 | 0.4281 | 0.2366 | 0.0704 |
| rs11125759 | 0.3718 | 0.2914 | 0.2020 |
| rs11263940 | 0.3203 | 0.2901 | 0.2694 |
| rs11768238 | 0.2408 | 0.3022 | 0.4257 |
| rs1555466 | 0.0501 | 0.2950 | 0.8650 |
| rs203768 | 0.2304 | 0.3066 | 0.4524 |
| rs2198140 | -0.0884 | 0.2769 | 0.7495 |
| rs2564930 | 0.7096 | 0.2623 | 0.0068 |
| rs34320 | -0.2928 | 0.2903 | 0.3132 |
| rs3899258 | 0.1694 | 0.2782 | 0.5425 |
| rs4129585 | 0.5476 | 0.2624 | 0.0369 |
| rs4702 | -0.1034 | 0.3005 | 0.7307 |
| rs4831130 | 0.3205 | 0.3182 | 0.3137 |
| rs4931 | -0.0851 | 0.3233 | 0.7924 |
| rs6474628 | 0.3951 | 0.3118 | 0.2051 |
| rs6660196 | 0.1518 | 0.3038 | 0.6172 |
| rs67839857 | 0.1522 | 0.3019 | 0.6143 |
| rs7626445 | 0.0348 | 0.2615 | 0.8940 |
| rs78587207 | 0.3225 | 0.2531 | 0.2025 |
| rs8003822 | 1.1372 | 0.2926 | 1.02e-04 |
| rs9479138 | 0.0796 | 0.3162 | 0.8013 |
| rs9886111 | -0.1733 | 0.3098 | 0.5759 |
| All - Inverse variance weighted | 0.2392 | 0.0678 | 4.18e-04 |
| All - MR Egger | 0.6114 | 0.7906 | 0.4484 |

**Scatter plot**

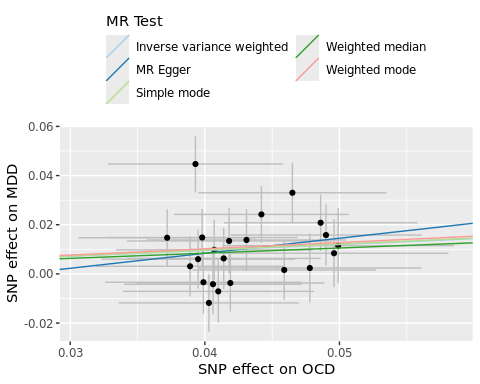

**Forest plot**

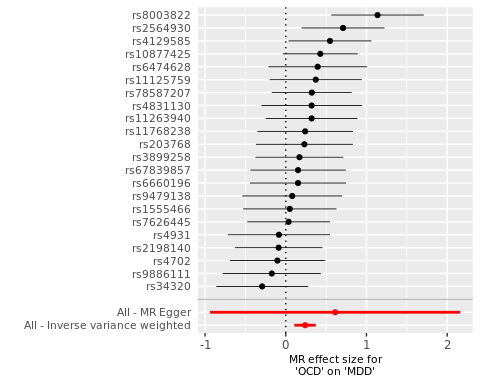

**Funnel plot**

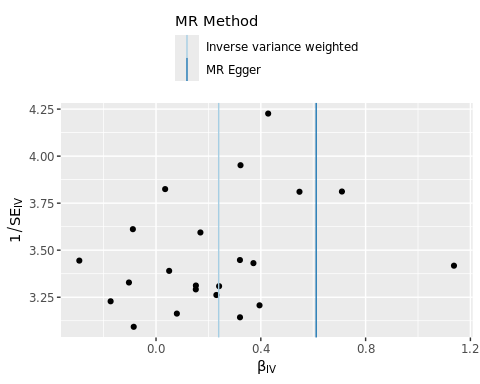

### BD-PGC3 → OCD

Table 8. MR-PRESSO Single SNP analysis results for BD-PGC3 → OCD

| SNP | b | se | p |
| --- | --- | --- | --- |
| rs10994415 | 0.1456 | 0.1008 | 0.1484 |
| rs113779084 | -0.0675 | 0.1033 | 0.5135 |
| rs12289486 | 0.0593 | 0.1283 | 0.6440 |
| rs13044225 | 0.1828 | 0.1243 | 0.1414 |
| rs1487445 | -0.0701 | 0.0876 | 0.4234 |
| rs1998820 | 0.0084 | 0.1266 | 0.9474 |
| rs2011302 | -0.0775 | 0.1304 | 0.5522 |
| rs2273738 | -0.1505 | 0.1101 | 0.1717 |
| rs2345964 | 0.1309 | 0.1218 | 0.2824 |
| rs237460 | 0.0542 | 0.1193 | 0.6499 |
| rs28455634 | -0.2166 | 0.1131 | 0.0554 |
| rs28565152 | 0.0134 | 0.1162 | 0.9081 |
| rs2989473 | 0.1443 | 0.1176 | 0.2199 |
| rs4331993 | -0.0365 | 0.1280 | 0.7753 |
| rs4672 | 0.0344 | 0.1190 | 0.7726 |
| rs475805 | -0.0045 | 0.1224 | 0.9708 |
| rs62489493 | 0.0937 | 0.1070 | 0.3813 |
| rs6456095 | 0.1815 | 0.1270 | 0.1531 |
| rs6865469 | -0.0721 | 0.1340 | 0.5904 |
| rs6954854 | 0.1423 | 0.1201 | 0.2360 |
| rs696366 | 0.2654 | 0.1376 | 0.0537 |
| rs7201930 | -0.0188 | 0.1194 | 0.8752 |
| rs72809838 | 0.1570 | 0.1088 | 0.1488 |
| All - Inverse variance weighted | 0.0331 | 0.0248 | 0.1818 |
| All - MR Egger | -0.0619 | 0.1191 | 0.6087 |

**Scatter plot**

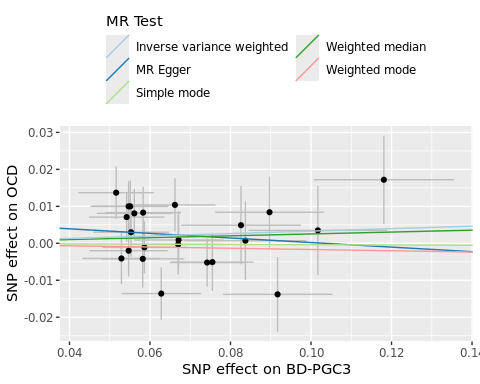

**Forest plot**

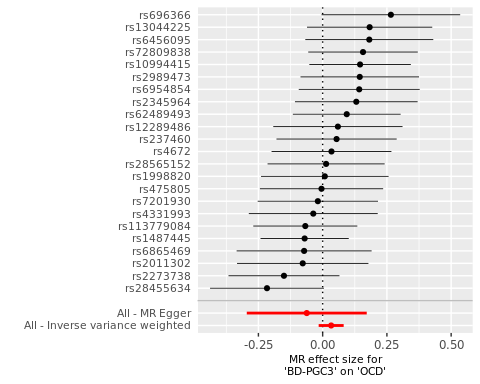

**Funnel plot**

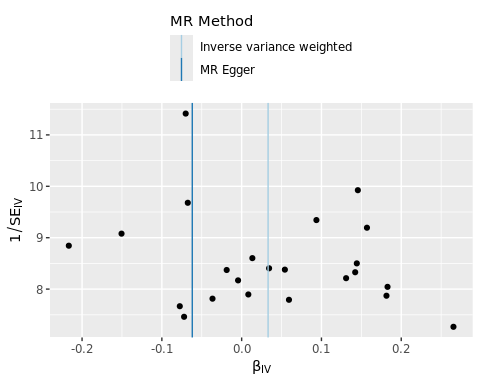

### BD1-PGC3 → OCD

Table 9. MR-PRESSO Single SNP analysis results for BD1-PGC3 → OCD

| SNP | b | se | p |
| --- | --- | --- | --- |
| rs10043984 | -0.2896 | 0.0928 | 0.0018 |
| rs10100074 | -0.0658 | 0.1024 | 0.5207 |
| rs10917509 | 0.0270 | 0.0974 | 0.7814 |
| rs11720159 | 0.2951 | 0.0946 | 0.0018 |
| rs11744542 | -0.0542 | 0.1057 | 0.6079 |
| rs12575685 | 0.0923 | 0.0899 | 0.3045 |
| rs1452386 | 0.0170 | 0.1017 | 0.8676 |
| rs1510606 | 0.2341 | 0.1052 | 0.0260 |
| rs174578 | 0.1485 | 0.0896 | 0.0976 |
| rs1924817 | -0.1336 | 0.1140 | 0.2411 |
| rs2314398 | 0.0953 | 0.1034 | 0.3567 |
| rs237475 | 0.0337 | 0.0855 | 0.6932 |
| rs2388334 | -0.0648 | 0.0844 | 0.4421 |
| rs28565152 | 0.0104 | 0.0898 | 0.9081 |
| rs3803277 | 0.0184 | 0.1011 | 0.8558 |
| rs4676412 | 0.0219 | 0.1028 | 0.8316 |
| rs4722033 | 0.0687 | 0.1061 | 0.5171 |
| rs4748468 | 0.0255 | 0.1003 | 0.7992 |
| rs489337 | -0.0138 | 0.0929 | 0.8822 |
| rs59127720 | -0.3688 | 0.1037 | 3.75e-04 |
| rs6073621 | 0.0130 | 0.0926 | 0.8879 |
| rs61074241 | 0.2651 | 0.0899 | 0.0032 |
| rs61846516 | 0.1986 | 0.0979 | 0.0425 |
| rs6495988 | 0.2095 | 0.0923 | 0.0233 |
| rs6993953 | -0.1500 | 0.0859 | 0.0808 |
| rs7248205 | -0.0252 | 0.1022 | 0.8052 |
| rs735931 | 0.0889 | 0.0993 | 0.3707 |
| rs9295354 | 0.1596 | 0.0989 | 0.1066 |
| rs9831123 | -0.3270 | 0.0959 | 6.51e-04 |
| rs9834970 | 0.1416 | 0.0626 | 0.0238 |
| All - Inverse variance weighted | 0.0292 | 0.0292 | 0.3175 |
| All - MR Egger | 0.2419 | 0.1903 | 0.2142 |

**Scatter plot**

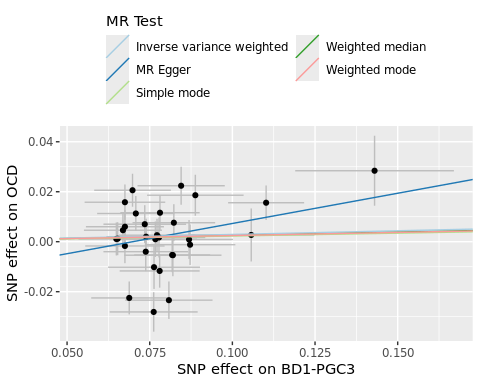

**Forest plot**

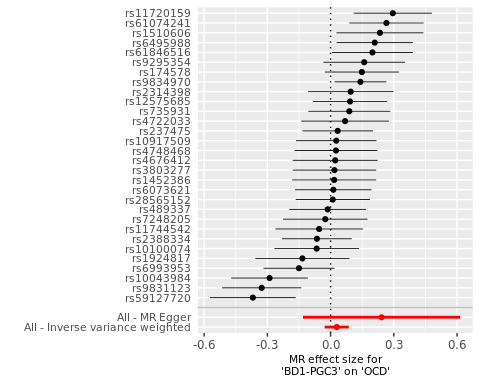

**Funnel plot**

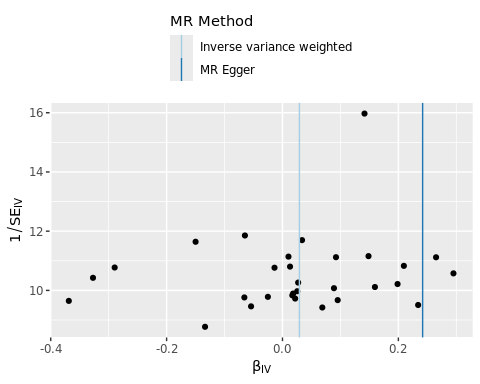

### BD-clinical-PGC4 → OCD

Table 10. MR-PRESSO Single SNP analysis results for BD-clinical-PGC4 → OCD

| SNP | b | se | p |
| --- | --- | --- | --- |
| rs10994318 | 0.2046 | 0.1030 | 0.0471 |
| rs11062170 | -0.1303 | 0.0807 | 0.1066 |
| rs112481526 | 0.2061 | 0.1130 | 0.0681 |
| rs112509803 | -0.0179 | 0.1044 | 0.8640 |
| rs113779084 | -0.0610 | 0.0934 | 0.5135 |
| rs1571617 | -0.1452 | 0.0920 | 0.1146 |
| rs2015582 | 0.1930 | 0.1158 | 0.0956 |
| rs2072262 | -0.0622 | 0.1110 | 0.5750 |
| rs2314398 | 0.0883 | 0.0958 | 0.3567 |
| rs237475 | 0.0391 | 0.0991 | 0.6932 |
| rs2388334 | -0.0717 | 0.0932 | 0.4421 |
| rs4331993 | -0.0336 | 0.1178 | 0.7753 |
| rs4676412 | 0.0248 | 0.1168 | 0.8316 |
| rs489337 | -0.0153 | 0.1031 | 0.8822 |
| rs496946 | 0.2139 | 0.1104 | 0.0526 |
| rs6062198 | 0.1509 | 0.1158 | 0.1924 |
| rs61554907 | 0.0009 | 0.1020 | 0.9928 |
| rs6558405 | -0.1818 | 0.0997 | 0.0682 |
| rs696366 | 0.2091 | 0.1084 | 0.0537 |
| rs7199910 | 0.0746 | 0.1183 | 0.5286 |
| rs7248205 | -0.0286 | 0.1160 | 0.8052 |
| rs7408675 | 0.2579 | 0.1215 | 0.0338 |
| rs7651996 | -0.3733 | 0.1100 | 6.89e-04 |
| rs814424 | 0.0099 | 0.0981 | 0.9194 |
| All - Inverse variance weighted | 0.0098 | 0.0302 | 0.7470 |
| All - MR Egger | -0.1167 | 0.1506 | 0.4466 |

**Scatter plot**

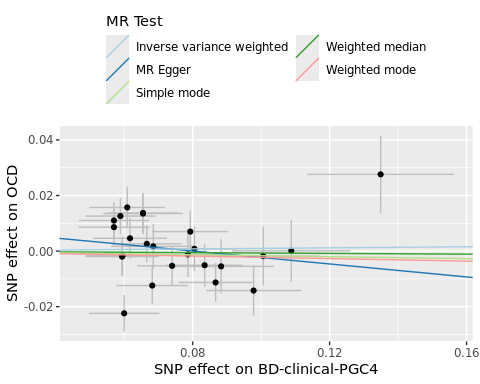

**Forest plot**

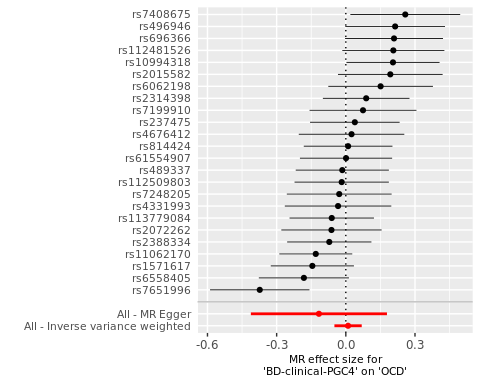

**Funnel plot**

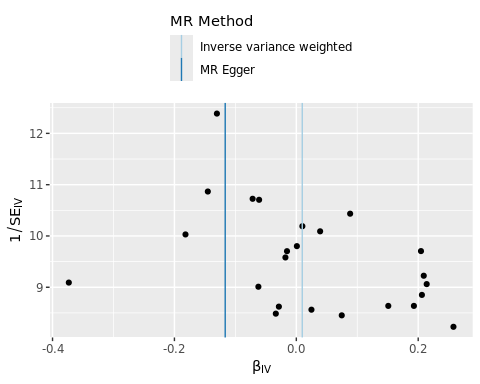

### SCZ → OCD

Table 11. MR-PRESSO Single SNP analysis results for SCZ → OCD

| SNP | b | se | p |
| --- | --- | --- | --- |
| rs10861176 | 0.0234 | 0.1423 | 0.8692 |
| rs10876446 | -0.0444 | 0.1426 | 0.7556 |
| rs11210892 | -0.0363 | 0.1087 | 0.7386 |
| rs11263861 | -0.0403 | 0.1363 | 0.7676 |
| rs11587347 | 0.2050 | 0.1155 | 0.0759 |
| rs12293670 | 0.0567 | 0.0979 | 0.5623 |
| rs12652777 | 0.1537 | 0.1352 | 0.2557 |
| rs13016542 | 0.1292 | 0.1098 | 0.2397 |
| rs16960158 | 0.1633 | 0.1411 | 0.2470 |
| rs1716180 | -0.2710 | 0.1030 | 0.0085 |
| rs1860002 | -0.0036 | 0.0847 | 0.9663 |
| rs187557 | 0.1319 | 0.1379 | 0.3389 |
| rs1901512 | 0.0120 | 0.1250 | 0.9236 |
| rs2333321 | 0.1602 | 0.1138 | 0.1591 |
| rs2455415 | -0.0283 | 0.1333 | 0.8319 |
| rs2532240 | 0.1036 | 0.1217 | 0.3946 |
| rs2909457 | -0.1286 | 0.1347 | 0.3398 |
| rs4630706 | 0.2213 | 0.1502 | 0.1407 |
| rs4936215 | 0.1603 | 0.1065 | 0.1323 |
| rs498591 | 0.1131 | 0.1283 | 0.3781 |
| rs56205728 | -0.2063 | 0.1190 | 0.0832 |
| rs56335113 | 0.0449 | 0.1113 | 0.6867 |
| rs58120505 | -0.1250 | 0.0737 | 0.0896 |
| rs61405217 | -0.0842 | 0.1383 | 0.5426 |
| rs634940 | 0.1733 | 0.1130 | 0.1251 |
| rs6546857 | 0.1474 | 0.1275 | 0.2477 |
| rs6943762 | 0.1798 | 0.0942 | 0.0562 |
| rs6984242 | -0.2578 | 0.1225 | 0.0354 |
| rs704367 | -0.1528 | 0.1154 | 0.1857 |
| rs7191183 | -0.0171 | 0.1301 | 0.8954 |
| rs72802868 | 0.0607 | 0.1040 | 0.5598 |
| rs7403630 | 0.1125 | 0.1343 | 0.4022 |
| rs79210963 | -0.0257 | 0.1238 | 0.8358 |
| rs79212538 | 0.2239 | 0.1439 | 0.1197 |
| rs893949 | 0.1260 | 0.1080 | 0.2434 |
| rs9454727 | 0.0975 | 0.1452 | 0.5020 |
| rs9636107 | 0.0987 | 0.0930 | 0.2887 |
| All - Inverse variance weighted | 0.0320 | 0.0220 | 0.1461 |
| All - MR Egger | 0.1667 | 0.1059 | 0.1245 |

**Scatter plot**

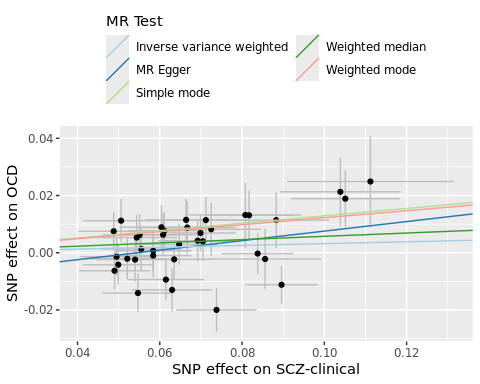

**Forest plot**

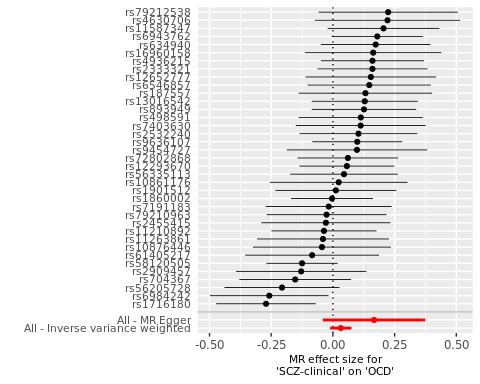

**Funnel plot**

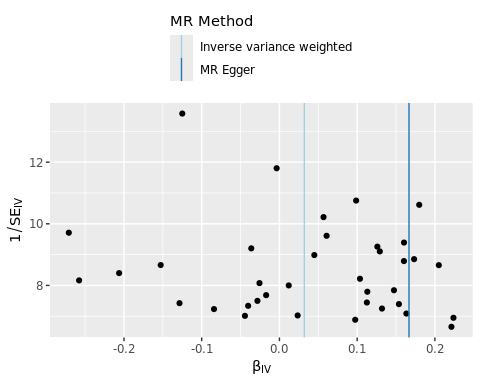

### OCD → BD1-PGC3 (removing borderline SNPs)

Table 12. MR-PRESSO Single SNP analysis results for OCD → BD1-PGC3 (removing borderline SNPs)

| SNP | b | se | p |
| --- | --- | --- | --- |
| rs10877425 | -0.0226 | 0.2448 | 0.9264 |
| rs11125759 | 0.1507 | 0.2914 | 0.6051 |
| rs11263940 | 0.7936 | 0.2901 | 0.0062 |
| rs11768238 | -0.0885 | 0.3022 | 0.7695 |
| rs1555466 | 0.0543 | 0.2908 | 0.8518 |
| rs203767 | 1.0701 | 0.3106 | 5.71e-04 |
| rs2087319 | -0.3790 | 0.3116 | 0.2239 |
| rs2198140 | 0.2577 | 0.2721 | 0.3435 |
| rs34320 | 0.0323 | 0.2903 | 0.9115 |
| rs3899258 | 0.0242 | 0.2802 | 0.9312 |
| rs4129585 | 0.4320 | 0.2624 | 0.0997 |
| rs4831130 | -0.2129 | 0.3182 | 0.5034 |
| rs4931 | 0.5190 | 0.3233 | 0.1085 |
| rs6474628 | 0.7875 | 0.3145 | 0.0123 |
| rs6660196 | -0.5190 | 0.3038 | 0.0876 |
| rs67839857 | -0.5339 | 0.3019 | 0.0770 |
| rs7626445 | 0.2222 | 0.2615 | 0.3954 |
| rs78587207 | -0.0020 | 0.2531 | 0.9936 |
| rs8003822 | 0.0025 | 0.2926 | 0.9931 |
| rs9479138 | 0.7068 | 0.3136 | 0.0242 |
| rs9886111 | -0.1999 | 0.3073 | 0.5154 |
| All - Inverse variance weighted | 0.1429 | 0.0908 | 0.1155 |
| All - MR Egger | 0.2851 | 1.0738 | 0.7935 |

**Scatter plot**

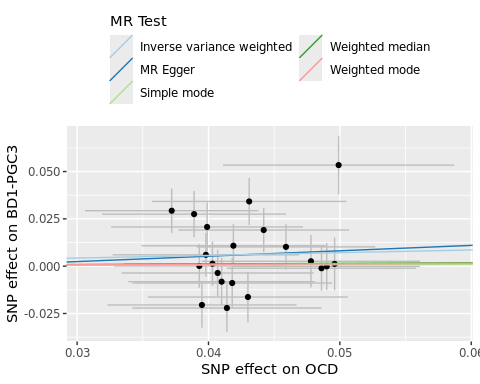

**Forest plot**

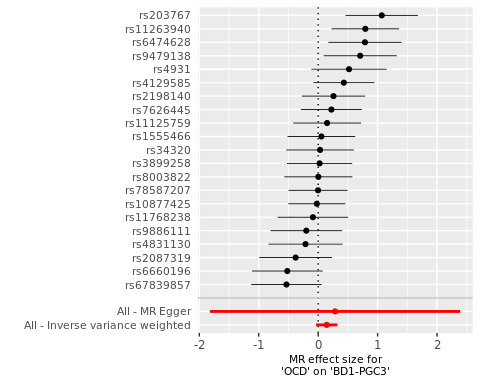

**Funnel plot**

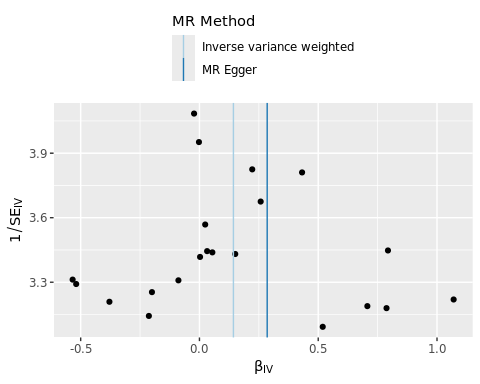

### BD1-PGC3 (removing borderline SNPs) → OCD

Table 13. MR-PRESSO Single SNP analysis results for BD1-PGC3 (removing borderline SNPs) → OCD

| SNP | b | se | p |
| --- | --- | --- | --- |
| rs10043984 | -0.2896 | 0.0928 | 0.0018 |
| rs10100074 | -0.0658 | 0.1024 | 0.5207 |
| rs10917509 | 0.0270 | 0.0974 | 0.7814 |
| rs11720159 | 0.2951 | 0.0946 | 0.0018 |
| rs11744542 | -0.0542 | 0.1057 | 0.6079 |
| rs12575685 | 0.0923 | 0.0899 | 0.3045 |
| rs1452386 | 0.0170 | 0.1017 | 0.8676 |
| rs1510606 | 0.2341 | 0.1052 | 0.0260 |
| rs174578 | 0.1485 | 0.0896 | 0.0976 |
| rs1924817 | -0.1336 | 0.1140 | 0.2411 |
| rs2314398 | 0.0953 | 0.1034 | 0.3567 |
| rs237475 | 0.0337 | 0.0855 | 0.6932 |
| rs2388334 | -0.0648 | 0.0844 | 0.4421 |
| rs28565152 | 0.0104 | 0.0898 | 0.9081 |
| rs3803277 | 0.0184 | 0.1011 | 0.8558 |
| rs4676412 | 0.0219 | 0.1028 | 0.8316 |
| rs4722033 | 0.0687 | 0.1061 | 0.5171 |
| rs4748468 | 0.0255 | 0.1003 | 0.7992 |
| rs489337 | -0.0138 | 0.0929 | 0.8822 |
| rs59127720 | -0.3688 | 0.1037 | 3.75e-04 |
| rs6073621 | 0.0130 | 0.0926 | 0.8879 |
| rs61074241 | 0.2651 | 0.0899 | 0.0032 |
| rs61846516 | 0.1986 | 0.0979 | 0.0425 |
| rs6495988 | 0.2095 | 0.0923 | 0.0233 |
| rs6993953 | -0.1500 | 0.0859 | 0.0808 |
| rs7248205 | -0.0252 | 0.1022 | 0.8052 |
| rs735931 | 0.0889 | 0.0993 | 0.3707 |
| rs9295354 | 0.1596 | 0.0989 | 0.1066 |
| rs9831123 | -0.3270 | 0.0959 | 6.51e-04 |
| rs9834970 | 0.1416 | 0.0626 | 0.0238 |
| All - Inverse variance weighted | 0.0292 | 0.0292 | 0.3175 |
| All - MR Egger | 0.2419 | 0.1903 | 0.2142 |

**Scatter plot**

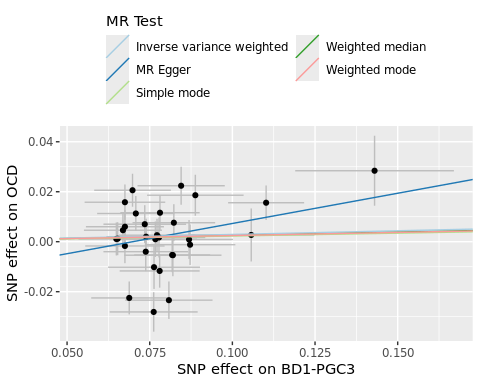

**Forest plot**

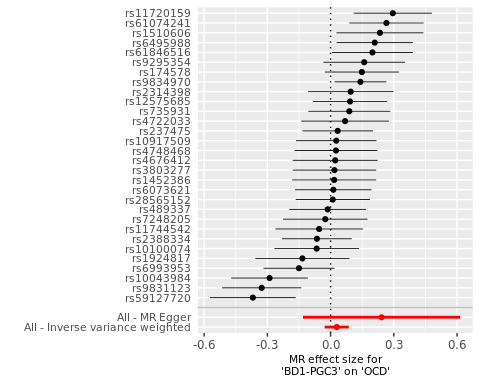

**Funnel plot**

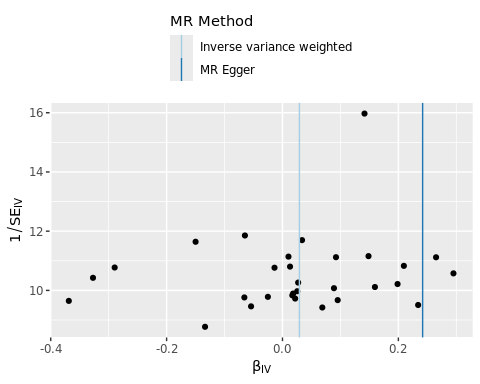
