## Supplementary material for "Directional genetic relationships between obsessive-compulsive disorder and bipolar disorder and schizophrenia": Strobe_Checklist

**STROBE-MR checklist of recommended items to address in reports of Mendelian randomization studies**^1^ ^2^

| **Item No.** | **Section** | **Checklist item** | **Page No.** | **Relevant text from manuscript** |
| --- | --- | --- | --- | --- |
| 1 | **TITLE and ABSTRACT** | Indicate Mendelian randomization (MR) as the study’s design in the title and/or the abstract if that is a main purpose of the study | Abstract | Using linkage disequilibrium score regression (LDSC), MiXeR, and Generalized Summary-data-based Mendelian Randomization (GSMR) as well as complementary Mendelian randomization approaches, we characterized genetic correlations, polygenic overlap (Dice coefficient), and effect direction concordance (ρβ) across disorders. |
|  | **INTRODUCTION** |  |  |  |
| 2 | **Background** | Explain the scientific background and rationale for the reported study. What is the exposure? Is a potential causal relationship between exposure and outcome plausible? Justify why MR is a helpful method to address the study question | 2 | “Two non-mutually exclusive explanations are: (i) shared genetic risk (pleiotropy)… or (ii) causal relationships between OCD and these psychiatric disorders”.  “Distinguishing among these possibilities is essential for clarifying whether observed cross-disorder associations reflect shared etiology, developmental progression, or diagnostic structure”.  “Here, we leverage the latest GWAS summary statistics… to systematically characterize their genetic correlation, polygenic overlap, and directional relationships using a suite of robust MR Methods”. |
| 3 | **Objectives** | State specific objectives clearly, including pre-specified causal hypotheses (if any). State that MR is a method that, under specific assumptions, intends to estimate causal effects | 2 | “Here, we leverage the latest GWAS summary statistics for OCD, BD (and its subtypes), SCZ, and MDD to systematically characterize their genetic correlation, polygenic overlap, and directional relationships…”  “…we aim to clarify whether OCD’s genetic relationships with BD, SCZ, and MDD reflect shared architecture, directional effects, or both, with implications for prognosis, early detection, and treatment stratification.” |
|  | **METHODS** |  |  |  |
| 4 | **Study design and data sources** | Present key elements of the study design early in the article. Consider including a table listing sources of data for all phases of the study. For each data source contributing to the analysis, describe the following: |  |  |
|  | a) | Setting: Describe the study design and the underlying population, if possible. Describe the setting, locations, and relevant dates, including periods of recruitment, exposure, follow-up, and data collection, when available. | 3 | “We used publicly available GWAS summary statistics from large-scale psychiatric consortia (Table 1). These datasets include the most recent and well-powered studies of OCD, BD, and its subtypes, SCZ, and MDD.”  “All analyses were constrained to individuals of European ancestry, as available GWAS summary statistics were predominantly derived from European-ancestry samples…”  “Full details of case definitions, control selection, and diagnostic assessment methods are provided in the original GWAS publications cited in Table 1.” |
|  | b) | Participants: Give the eligibility criteria, and the sources and methods of selection of participants. Report the sample size, and whether any power or sample size calculations were carried out prior to the main analysis | 4-6 | “We used publicly available GWAS summary statistics from large-scale psychiatric consortia…”  “For each exposure and outcome, disease status was defined in the source GWAS using standardized diagnostic criteria… Full details of case definitions, control selection, and diagnostic assessment methods are provided in the original GWAS publications…”  “OCD cases were defined using clinician-confirmed diagnoses and/or structured diagnostic interviews aligned with DSM-IV or DSM-5 criteria… Control participants were screened to exclude OCD and other major psychiatric disorders when possible.”  “Power calculations indicated that we had 80% power to detect small effect sizes (OR 1.02 to 1.14) across all analyses…” |
|  | c) | Describe measurement, quality control and selection of genetic variants | 5-6 | “Genetic variants were measured as single nucleotide polymorphisms (SNPs) represented by per-allele effect size estimates and standard errors from GWAS summary statistics.”  “All variants included in the present analyses had undergone quality control, including standard procedures for genotyping, imputation, and variant-level filtering, as described in the respective source publications.”  “Missing genotype or phenotype data were addressed in the original GWAS through study-specific quality control and imputation procedures…”  “In the present analyses, only SNPs with complete summary statistics (effect size estimates and standard errors) available for both exposure and outcome datasets were included.”  “Genome-wide significant SNPs (p < 5×10⁻⁸) were selected as instrumental variables (IVs) and pruned for LD (r² < 0.01) based on the 1000 Genomes European reference panel.”  “To mitigate bias for horizontal pleiotropy, the HEIDI-outlier method (p < 0.01) was used to identify and exclude pleiotropic variants.” |
|  | d) | For each exposure, outcome, and other relevant variables, describe methods of assessment and diagnostic criteria for diseases | 4-5 | “For each exposure and outcome, disease status was defined in the source GWAS using standardized diagnostic criteria appropriate to each disorder…”  “OCD cases were defined using clinician-confirmed diagnoses and/or structured diagnostic interviews aligned with DSM-IV or DSM-5 criteria. In addition, large population-based cohorts… contributed self-reported OCD diagnoses. Control participants were screened to exclude OCD and other major psychiatric disorders when possible.”  “BD cases were diagnosed using standardized criteria consistent with DSM-IV, based on structured interviews, clinician assessment, or medical record review… BD1… defined by the presence of manic episodes, whereas BD2… required hypomanic episodes and major depressive episodes without a history of full mania.”  “Schizophrenia cases were diagnosed according to DSM-IV or ICD-10 criteria using structured clinical interviews, clinician assessment, or medical record review. Controls were screened to exclude psychotic disorders.”  “MDD cases were defined… using structured diagnostic interviews, clinician diagnosis, or validated questionnaire-based assessments consistent with DSM-IV or DSM-5 criteria. Control participants were screened to exclude lifetime MDD diagnosis.” |
|  | e) | Provide details of ethics committee approval and participant informed consent, if relevant | 5 | “Ethical approval for each contributing GWAS was obtained from the relevant institutional review boards in the original studies, and our analysis was not pre-registered. The present study used de-identified summary data and did not require additional ethical approval” |
| 5 | **Assumptions** | Explicitly state the three core IV assumptions for the main analysis (relevance, independence and exclusion restriction) as well assumptions for any additional or sensitivity analysis | 5 | “Mendelian randomization uses genetic variants as instrumental variables to estimate the effect of an exposure on an outcome…”  “Causal inference using MR relies on three core assumptions: (i) the genetic instruments are robustly associated with the exposure (relevance); (ii) the instruments are independent of confounders of the exposure–outcome relationship (independence); and (iii) the instruments influence the outcome only through the exposure and not through alternative biological pathways (exclusion restriction).”  “Violations of the exclusion restriction assumption may occur in the presence of horizontal pleiotropy, whereby genetic variants affect the outcome through pathways independent of the exposure.”  “To evaluate the robustness of findings… we conducted complementary analyses using the TwoSampleMR package… including inverse-variance weighted (IVW), MR-Egger, weighted median, simple mode, and weighted mode estimators.”  “Between-instrument heterogeneity was quantified using Cochran’s Q… Directional horizontal pleiotropy was assessed using the MR-Egger intercept test… outlier-driven pleiotropy was evaluated using MR-PRESSO…” |
| 6 | **Statistical methods: main analysis** | Describe statistical methods and statistics used |  |  |
|  | a) | Describe how quantitative variables were handled in the analyses (i.e., scale, units, model) | 5-6 | “All traits were binary, and SNP effects were modelled on the log-odds scale.”  “Accordingly, causal estimates reflect the change in log-odds of the outcome per one-unit increase in genetically predicted liability to the exposure.”  “No additional transformations were applied.” |
|  | b) | Describe how genetic variants were handled in the analyses and, if applicable, how their weights were selected | 5-6 | “Genetic variants were measured as single nucleotide polymorphisms (SNPs) represented by per-allele effect size estimates and standard errors from GWAS summary statistics.”  “In the present analyses, only SNPs with complete summary statistics (effect size estimates and standard errors) available for both exposure and outcome datasets were included.”  “Genome-wide significant SNPs (p < 5×10⁻⁸) were selected as instrumental variables (IVs) and pruned for LD (r² < 0.01) based on the 1000 Genomes European reference panel.”  “To mitigate bias for horizontal pleiotropy, the HEIDI-outlier method (p < 0.01) was used to identify and exclude pleiotropic variants.”  “These sensitivity analyses included inverse-variance weighted (IVW), MR-Egger, weighted median, simple mode, and weighted mode estimators.” |
|  | c) | Describe the MR estimator (e.g. two-stage least squares, Wald ratio) and related statistics. Detail the included covariates and, in case of two-sample MR, whether the same covariate set was used for adjustment in the two samples | 5-6 | “We employed Generalised Summary-data-based Mendelian Randomisation (GSMR)… to investigate potential bidirectional causal relationships…”  “Genome-wide significant SNPs… were selected as instrumental variables (IVs)…”  “To evaluate the robustness of findings… we conducted complementary analyses using the TwoSampleMR package… These sensitivity analyses included inverse-variance weighted (IVW), MR-Egger, weighted median, simple mode, and weighted mode estimators.”  “Between-instrument heterogeneity was quantified using Cochran’s Q statistics…”  “Directional horizontal pleiotropy was assessed using the MR-Egger intercept test, and outlier-driven pleiotropy was evaluated using MR-PRESSO.”  “Instrument strength was evaluated using the mean F-statistic…”  “Covariate adjustment (e.g., age, sex, ancestry principal components) was performed within the original GWAS from which summary statistics were derived.” |
|  | d) | Explain how missing data were addressed | 5 | “Missing genotype or phenotype data were addressed in the original GWAS through study-specific quality control and imputation procedures, as described in the respective source publications.”  “In the present analyses, only SNPs with complete summary statistics (effect size estimates and standard errors) available for both exposure and outcome datasets were included.” |
|  | e) | If applicable, indicate how multiple testing was addressed | 6 | “We did not apply multiple testing correction, as GSMR was specified as the primary analysis. Complementary Mendelian randomization methods were used as sensitivity analyses to assess robustness of findings rather than as independent hypothesis tests”. |
| 7 | **Assessment of assumptions** | Describe any methods or prior knowledge used to assess the assumptions or justify their validity | 5-6 | “Violations of the exclusion restriction assumption may occur in the presence of horizontal pleiotropy…”  “To mitigate bias for horizontal pleiotropy, the HEIDI-outlier method (p < 0.01) was used to identify and exclude pleiotropic variants.”  “Between-instrument heterogeneity was quantified using Cochran’s Q statistics…”  “Directional horizontal pleiotropy was assessed using the MR-Egger intercept test, and outlier-driven pleiotropy was evaluated using MR-PRESSO.”  “To assess the reliability of MR-Egger estimates, we calculated the I²GX statistic as a measure of potential regression dilution bias.”  “Directionality was further assessed using Steiger filtering…”  “Concordance in the direction and magnitude of effect estimates across methods was used to support the stability of GSMR findings.” |
| 8 | **Sensitivity analyses and additional analyses** | Describe any sensitivity analyses or additional analyses performed (e.g. comparison of effect estimates from different approaches, independent replication, bias analytic techniques, validation of instruments, simulations) | 5 | “To evaluate the robustness of findings and further explore potential violations of these assumptions, we conducted complementary analyses using the TwoSampleMR package…”  “These sensitivity analyses included inverse-variance weighted (IVW), MR-Egger, weighted median, simple mode, and weighted mode estimators.”  “Between-instrument heterogeneity was quantified using Cochran’s Q statistics…”  “Directional horizontal pleiotropy was assessed using the MR-Egger intercept test, and outlier-driven pleiotropy was evaluated using MR-PRESSO.”  “Instrument strength was evaluated using the mean F-statistic…” |
| 9 | **Software and pre-registration** |  |  |  |
|  | a) | Name statistical software and package(s), including version and settings used |  | “We applied MiXeR (v1.3) to all OCD-disorder pairs.”  “We employed Generalised Summary-data-based Mendelian Randomisation (GSMR) implemented in GCTA software (version 1.94.1)…”  “We conducted complementary analyses using the TwoSampleMR package (v0.5.6) in R.” |
|  | b) | State whether the study protocol and details were pre-registered (as well as when and where) | 5 | “Our analysis was not pre-registered.” |
|  | **RESULTS** |  |  |  |
| 10 | **Descriptive data** |  |  |  |
|  | a) | Report the numbers of individuals at each stage of included studies and reasons for exclusion. Consider use of a flow diagram | Table 1 | Table 1 reports numbers of cases and controls for each dataset) |
|  | b) | Report summary statistics for phenotypic exposure(s), outcome(s), and other relevant variables (e.g. means, SDs, proportions) | n/a | Not applicable; individual-level phenotypic data were not available. Phenotypic summaries are reported in the source GWAS publications. |
|  | c) | If the data sources include meta-analyses of previous studies, provide the assessments of heterogeneity across these studies | n/a |  |
|  | d) | For two-sample MR:  i.  Provide justification of the similarity of the genetic variant-exposure associations between the exposure and outcome samples  ii.  Provide information on the number of individuals who overlap between the exposure and outcome studies | 4  6 | “All analyses were constrained to individuals of European ancestry, as available GWAS summary statistics were predominantly derived from European-ancestry samples; this restriction also minimized confounding due to population structure.”  “Information regarding participant-level overlap between exposure and outcome GWAS was not available.”  “Although partial sample overlap across psychiatric consortia cannot be ruled out, any such overlap is expected to represent a small fraction of the total sample size and is therefore unlikely to substantially bias effect estimates.” |
| 11 | **Main results** |  |  |  |
|  | a) | Report the associations between genetic variant and exposure, and between genetic variant and outcome, preferably on an interpretable scale | 7 | “Individual SNP associations are summarized through aggregate MR estimates, with full results provided in Table 4 and STables 2 to 5.”  “b denotes the GSMR effect estimate; s.e., standard error; NSNP, number of genetic instruments (instrumental variables).” |
|  | b) | Report MR estimates of the relationship between exposure and outcome, and the measures of uncertainty from the MR analysis, on an interpretable scale, such as odds ratio or relative risk per SD difference | 6 | “A one standard deviation increase in OCD genetic liability was associated with increased risk of BD across all definitions, including BD-PGC3 (b = 0.20, s.e. = 0.05, p = 1.5 × 10⁻⁴)… ”  “OCD liability was also strongly associated with SCZ (b = 0.52, s.e. = 0.06, p = 9.5 × 10⁻²¹) and with MDD (b = 0.24, s.e. = 0.06, p = 1.1 × 10⁻⁴).”  “b denotes the GSMR effect estimate; s.e., standard error…” |
|  | c) | If relevant, consider translating estimates of relative risk into absolute risk for a meaningful time period | n/a |  |
|  | d) | Consider plots to visualize results (e.g. forest plot, scatterplot of associations between genetic variants and outcome versus between genetic variants and exposure) |  | Supplement |
| 12 | **Assessment of assumptions** |  |  |  |
|  | a) | Report the assessment of the validity of the assumptions | 7-9 | “Although heterogeneity was observed for BD1-PGC3 and BD-clinical-PGC4, MR-Egger intercept tests and MR-PRESSO analyses did not indicate substantial directional pleiotropy, and exclusion of identified outliers did not materially alter the causal estimates.”  “Steiger filtering supported the hypothesized causal direction… with approximately 100% of variants explaining greater variance in the exposure than in OCD…”  “Cochran’s Q statistics indicated heterogeneity in most BD analyses, whereas MR-Egger intercept tests were generally non-significant…”  “MR-PRESSO identified evidence of horizontal pleiotropy… however, distortion tests were not significant, and exclusion of identified outliers did not substantially alter effect estimates.”  “For the effect of OCD on SCZ… MR-Egger intercept test showed no evidence of directional pleiotropy…”  “Steiger filtering provided limited support for the assumed causal direction…” |
|  | b) | Report any additional statistics (e.g., assessments of heterogeneity across genetic variants, such as *I^2^*, Q statistic or E-value) | 7-9 | “Cochran’s Q statistics indicated heterogeneity in most BD analyses…”  “Although heterogeneity was observed for BD1-PGC3 and BD-clinical-PGC4…”  “MR-Egger intercept tests were generally non-significant…”  “MR-PRESSO identified evidence of horizontal pleiotropy…” |
| 13 | **Sensitivity analyses and additional analyses** |  |  |  |
|  | a) | Report any sensitivity analyses to assess the robustness of the main results to violations of the assumptions | 7-9 | “When OCD was modeled as the outcome, sensitivity analyses using TwoSampleMR were broadly consistent with the primary GSMR findings…”  “Across BD phenotypes, effect estimates were small and non-significant across IVW and weighted median models…”  “Although heterogeneity was observed… MR-Egger intercept tests and MR-PRESSO analyses did not indicate substantial directional pleiotropy…”  “Exclusion of identified outliers did not materially alter the causal estimates.”  “When OCD was modeled as the exposure, sensitivity analyses were consistent with the GSMR findings across all outcomes…”  “IVW and weighted median estimates were directionally concordant with GSMR…”  “MR-PRESSO identified evidence of horizontal pleiotropy… however, distortion tests were not significant…”  “All MR methods yielded overlapping effect size estimates and consistent directions of effect…” |
|  | b) | Report results from other sensitivity analyses or additional analyses | 7-9 | “Power calculations indicated that we had 80% power to detect small effect sizes (OR 1.02 to 1.14) across all analyses…”  “Instrument strength criteria (mean F > 10)… supporting the reliability of the MR analyses.”  “Steiger filtering supported the hypothesized causal direction…”  “MR-PRESSO identified evidence of horizontal pleiotropy… however, distortion tests were not significant…”  “Exclusion of identified outliers did not substantially alter effect estimates…” |
|  | c) | Report any assessment of direction of causal relationship (e.g., bidirectional MR) | 6-8 | “We employed Generalised Summary-data-based Mendelian Randomisation (GSMR)… to investigate potential bidirectional causal relationships between genetic liability to OCD and BD, SCZ, and MDD.”  “We tested OCD → BD/SCZ/MDD and BD/SCZ/MDD → OCD directions.”  “Bidirectional GSMR analyses indicated asymmetric genetic relationships between OCD and the comparison disorders…”  “When OCD was modeled as the outcome… no evidence of causal effects…”  “In contrast, when OCD was modeled as the exposure, strong and consistent effects were observed on all outcomes…”  “Steiger filtering supported the hypothesized causal direction…” |
|  | d) | When relevant, report and compare with estimates from non-MR analyses | n/a |  |
|  | e) | Consider additional plots to visualize results (e.g., leave-one-out analyses) |  | Supplement |
|  | **DISCUSSION** |  |  |  |
| 14 | **Key results** | Summarize key results with reference to study objectives | 9-10 | “Our study provides the first comprehensive evaluation of shared genetic architecture and bidirectional causal relationships between OCD and other major psychiatric conditions, namely SCZ, BD, and MDD.”  “Beyond confirming substantial genetic correlations between OCD and each of these disorders… we quantify the degree of polygenic overlap and provide convergent evidence supporting directional genetic effects from OCD liability to BD, SCZ, and MDD.”  “Together, these findings refine our understanding of cross-disorder genetic architecture and support a model in which OCD liability represents an upstream component of transdiagnostic psychiatric risk.” |
| 15 | **Limitations** | Discuss limitations of the study, taking into account the validity of the IV assumptions, other sources of potential bias, and imprecision. Discuss both direction and magnitude of any potential bias and any efforts to address them | 11-12 | “These results, however, should also be interpreted cautiously given the reliance on summary-level data, the possibility of residual pleiotropy despite extensive sensitivity analyses, and limited power for some reverse-direction tests.”  “Causal interpretation in Mendelian randomization depends on the validity of underlying instrumental variable assumptions.”  “MR estimates rely on the assumption that genetic liability to a disorder operates through the same biological pathways as the disorder itself… this equivalence may be imperfect.”  “Evidence of heterogeneity across genetic instruments was observed… Such heterogeneity is not uncommon in MR studies of complex traits, and does not, in itself, imply directional horizontal pleiotropy.”  “All analyses were constrained to individuals of European ancestry… limits the generalizability…”  “The limited number of genome-wide significant instruments for some BD subtypes reduced statistical power for reverse-direction Mendelian randomization…”  “Partial sample overlap… could bias estimates toward observational associations; however… unlikely to substantially bias effect estimates.” |
| 16 | **Interpretation** |  |  |  |
|  | a) | Meaning: Give a cautious overall interpretation of results in the context of their limitations and in comparison with other studies | 9-11 | “Together, these findings refine our understanding of cross-disorder genetic architecture and support a model in which OCD liability represents an upstream component of transdiagnostic psychiatric risk.”  “These findings highlight both shared and disorder-specific components of the genetic architecture…”  “Collectively, these findings suggest that genetic liability to OCD is causally associated with liability to other major psychiatric disorders… however… findings are best interpreted as evidence of directional genetic influence rather than direct clinical causation…”  “While Mendelian randomization estimates reflect the effects of lifelong genetic liability rather than the consequences of clinical diagnosis or treatment…”  “These findings may have clinical relevance… however… should not be interpreted as evidence that intervention on OCD will prevent subsequent illness.”  “Consistent with prior work… we confirm substantial genetic correlations…” |
|  | b) | Mechanism: Discuss underlying biological mechanisms that could drive a potential causal relationship between the investigated exposure and the outcome, and whether the gene-environment equivalence assumption is reasonable. Use causal language carefully, clarifying that IV estimates may provide causal effects only under certain assumptions | 10-11 | “The observed directional genetic associations may reflect shared neurobiological pathways through which liability to OCD contributes to broader psychiatric vulnerability.”  “OCD has been linked to alterations in cortico-striatal-thalamo-cortical circuitry, glutamatergic and serotonergic signaling, and neurodevelopmental processes that overlap with pathways implicated in BD, SCZ, and MDD.”  “Emerging genetic and transcriptomic evidence suggests that OCD risk is enriched in genes involved in chromatin regulation, pointing to upstream mechanisms that influence gene expression during brain development…”  “This raises the possibility that genetic liability to OCD may influence early neurodevelopmental trajectories… increasing susceptibility to later-onset psychiatric conditions.”  “MR estimates rely on the assumption that genetic liability to a disorder operates through the same biological pathways as the disorder itself… this equivalence may be imperfect.”  “Accordingly, the present findings are best interpreted as evidence of directional genetic influence rather than direct clinical causation…” |
|  | c) | Clinical relevance: Discuss whether the results have clinical or public policy relevance, and to what extent they inform effect sizes of possible interventions | 11 | “These findings may have clinical relevance for understanding patterns of psychiatric comorbidity.”  “Clarifying the biology of these disorders may help determine whether comorbid presentations represent shared etiological mechanisms, developmental progression or distinct co-occurring conditions, which in turn has implications for treatment.”  “From a clinical perspective, this supports the importance of careful longitudinal assessment and monitoring for emerging mood or psychotic symptoms in individuals with OCD…”  “However, the present findings do not inform the expected magnitude of benefit from treating OCD on the risk of other psychiatric disorders, and should not be interpreted as evidence that intervention on OCD will prevent subsequent illness.” |
| 17 | **Generalizability** | Discuss the generalizability of the study results (a) to other populations, (b) across other exposure periods/timings, and (c) across other levels of exposure | 11-12 | “All analyses were constrained to individuals of European ancestry… This Eurocentric imbalance limits the generalizability of our findings to non-European populations…”  “MR estimates reflect effects on latent liability rather than clinical diagnosis.”  “MR estimates reflect the effects of lifelong genetic liability rather than the consequences of clinical diagnosis or treatment…” |
|  | **OTHER INFORMATION** |  |  |  |
| 18 | **Funding** | Describe sources of funding and the role of funders in the present study and, if applicable, sources of funding for the databases and original study or studies on which the present study is based | 1 | Funding is in title page. |
| 19 | **Data and data sharing** | Provide the data used to perform all analyses or report where and how the data can be accessed, and reference these sources in the article. Provide the statistical code needed to reproduce the results in the article, or report whether the code is publicly accessible and if so, where |  | “GWAS summary statistics used in this study are publicly available from the respective consortia as cited in Table 1. Statistical code used to perform the analyses is publicly available on GitHub at https://github.com/buxbaum-lab/OCD-BD” |
| 20 | **Conflicts of Interest** | All authors should declare all potential conflicts of interest | 1 | None reported. |

This checklist is copyrighted by the Equator Network under the Creative Commons Attribution 3.0 Unported (CC BY 3.0) license.

1.	Skrivankova VW, Richmond RC, Woolf BAR, Yarmolinsky J, Davies NM, Swanson SA, et al. Strengthening the Reporting of Observational Studies in Epidemiology using Mendelian Randomization (STROBE-MR) Statement. JAMA. 2021;under review.

2.	Skrivankova VW, Richmond RC, Woolf BAR, Davies NM, Swanson SA, VanderWeele TJ, et al. Strengthening the Reporting of Observational Studies in Epidemiology using Mendelian Randomisation (STROBE-MR): Explanation and Elaboration. BMJ. 2021;375:n2233.
